# PD-1/LAG-3 Dysfunctionality Signatures in Human Cancers

**DOI:** 10.1101/2023.03.10.23287087

**Authors:** Luisa Chocarro, Leticia Fernandez-Rubio, María Jesús García-Granda, Ester Blanco, Ana Bocanegra, Miriam Echaide, Maider Garnica, Miren Zuazo, Colette Johnston, Carolyn J. Edwards, James Legg, Andrew J Pierce, Hugo Arasanz, Ruth Vera, Karina Ausin, Enrique Santamaría, Joaquín Fernández-Irigoyen, Grazyna Kochan, David Escors

## Abstract

A significant number of cancer patients do not benefit from PD-L1/PD-1 blockade immunotherapies. PD-1 and LAG-3 co-upregulation in T-cells is one of the major mechanisms of resistance by establishing a highly dysfunctional state in T-cells. To identify shared features associated to PD- 1/LAG-3 dysfunctionality in human cancers and T-cells, multiomic expression profiles were obtained for all TCGA cancers with high T-cell infiltration. A PD-1/LAG-3 dysfunctional signature was found which regulated immune, metabolic, genetic and epigenetic pathways. These results were validated in T-cell lines with constitutively active PD-1, LAG-3 pathways and their combination. These results uncovered distinct degrees of T-cell dysfunctionality. Global changes were cross-evaluated among T-cell lines with multiomic biopsy data to identify genetic, metabolic, and proteomic programmes regulated by PD-1/LAG-3 dysfunctionality. One of these relied on differential regulation of E3 ubiquitin ligases CBL-B and C-CBL. PD-1/LAG-3 co-blockade with a bispecific drug under clinical development but not with a combination of anti-PD-1/anti-LAG-3 antibodies achieved both CBL-B and C-CBL inhibition, reverting T-cell dysfunctionality in lung cancer patients resistant to PD-L1/PD- 1 blockade.

## INTRODUCTION

PD-L1/PD-1 immune checkpoint (IC) blockade has revolutionised oncology (Topalian et al., 2015; Topalian et al., 2016). This strategy reactivates cancer-specific T-cells leading to tumour regression or long-term disease control. However, intrinsic resistance to PD-L1/PD-1 blockade frequently occurs through several mechanisms. One of the most relevant is the co-expression of LAG-3 with PD-1 in T- cells, stablishing strong T-cell dysfunctionality in patients resistant to conventional immunotherapies, as we and others have shown (Chocarro et al., 2022a; Zuazo et al., 2020; Zuazo et al., 2019). T-cell dysfunctionality can be partially overcome *ex vivo* with PD-1 and LAG-3 co-blockade in T-cells from non-small cell lung cancer (NSCLC) patients, which recover proliferative and effector activities (Edwards et al., 2021; Zuazo et al., 2019). These results indicate that PD-1 and LAG-3 cooperatively establish a strong dysfunctional estate in T-cells through co-signalling. The mechanisms by which PD- 1 interferes with T-cell functions are well-known and reviewed elsewhere (Arasanz et al., 2017). Briefly, PD-1 signalling terminates T-cell receptor (TCR) signal transduction, causes its down- modulation and degradation, and impacts on T-cell metabolism (Chemnitz et al., 2004; Hui et al., 2017; Karwacz et al., 2011; Patsoukis et al., 2015; Patsoukis et al., 2013; Plas et al., 1996; Sheppard et al., 2004). However, the specific inhibitory mechanisms regulated by LAG-3, and the combined effects of PD-1 and LAG-3 co-signalling are largely unknown (Edwards et al., 2021; Zuazo et al., 2019). It has been suggested that PD-1 and LAG-3 stablish a complex together with TCR components, that would be required for their combined inhibitory activities (Chen and Flies, 2013; Hannier et al., 1998; Iouzalen et al., 2001; Lichtenegger et al., 2018; Saito et al., 2010).

The clinical significance of PD-1/LAG-3 co-blockade has been recently demonstrated by the approval of Opdualag, a therapy combining anti-PD-1 (nivolumab) and anti-LAG-3 (relatlimab) antibodies. This strategy doubled survival of melanoma patients compared to PD-1 blockade alone (Chocarro et al., 2022a; Paik, 2022). However, it is unclear whether all tumour types may benefit from this combination, highlighting the importance of identifying common signatures of PD-1/LAG-3 immune dysfunctionality in T-cell infiltrated human cancers. The therapeutic activities of PD-1 and LAG-3 co- blockade may also differ when using single molecule bispecific blockers, as these agents possess stronger affinities towards PD-1^+^LAG-3^+^ T-cells. Some bispecific drugs have shown superior activities compared to combinations of anti-PD-1 with anti-LAG-3 agents (Chocarro et al., 2022b; Dahlen et al., 2018; Edwards et al., 2021; Jiang et al., 2021; Sung et al., 2022). Although much has been discovered on PD-1 functions and its blockade, the molecular effects of PD-1/LAG-3 co-blockade over PD- 1/LAG-3-associated dysfunctional signatures in cancer cells and T-cells are largely unknown.

Here we have systematically analysed extensive multiomic biopsy data through an initial selection based on high T-cell infiltration, and then extracting PD-1 and LAG-3 common omic signatures in all TCGA cancers. These signatures and their relationships with other immune checkpoint and tumour microenvironment molecules were established. PD-1/LAG-3-associated signatures were then studied in T-cell lines expressing PD-1 and LAG-3-based constructs that provided sustained signalling. Specific PD-1/LAG-3 co-signalling signatures brought to light an extensive and highly regulated programme associated to strong T-cell dysfunctionality in cancer. Two of the identified pathways were characterized following PD-1/LAG-3 co-blockade in primary dysfunctional T-cells from NSCLC patients by antibody combinations or by a bispecific molecule under clinical development, finding relevant molecular and functional differences.

## RESULTS

### PD-1/LAG-3 omic signatures in human cancers and association with other immune checkpoints

PD-1 together with LAG-3 establish a highly dysfunctional state in T-cells conferring resistance to PD-L1/PD-1 blockade (Edwards et al., 2021). To extend this finding to human tumours, we investigated whether dysfunctional gene signatures associated to PD-1/LAG-3 co-expression could be identified from publically available multi-omics biopsy data, and if they were related to immune infiltration, expression of additional IC molecules and activation of cancer cell pathways. Public transcriptomic and genomic databases for all TCGA human cancers were analysed with TIMER 2.0, Gene Expression Profiling Interactive Analysis (GEPIA), cBioportal, TNM Plot and Ingenuity Pathway Analysis (IPA). Overall, this study encompassed more than 12000 patients distributed into 40 cancer types.

First, a cBioportal-curated set of non-redundant multidimensional cancer genomics data (67603 case sets in the selected cohorts), which included TCGA and non-TCGA studies with no overlapping samples, confirmed that overall survival (OS) was reduced in patients with tumour signatures represented by two or more immune checkpoints **(Table S1)**. Then, *PDCD1*/*LAG3*-associated gene profiles were correlated with those associated to other immune checkpoints. To this end, gene expression of *CTLA4, TIM3, TIGIT, GITR, VISTA* and their associated gene profiles were analysed with TIMER 2.0 and IPA **(Figure 1)**. Thus, gene expression for each IC was quantified across all tumour samples together with paired normal tissue and compared between them **(Figure 1a)**. We found that median expression for each IC molecule differed among human cancers. Some specific cancers presented more elevated IC expression **(Figure 1a).** Then, pairwise correlations of IC expression were carried out **(Figure 1b)**. As expected, all immune checkpoints had positive correlations in most cancers. However, we found that *PDCD1, LAG3,* and *CTLA4* followed similar patterns across tumours, while *TIM3* and *TIGIT* differed. Overall, we also confirmed that IC overexpression also correlated with TOX, a critical regulator of T-cell exhaustion for most cancers with the notable exception of thymic cancer (THYM) **(Figure 1b, c)**. Then, we correlated IC expression with immune-related genes in all represented human cancers using global purity-adjusted partial Spearmańs correlation. Again, this analysis uncovered distinct gene expression profiles associated to *PDCD1/LAG3/CTLA4* or to *TIM3 /TIGIT*, while *GITR* and *VISTA* profiles were independent from the other ICs **(Figure 1d).**

**Figure 1.**
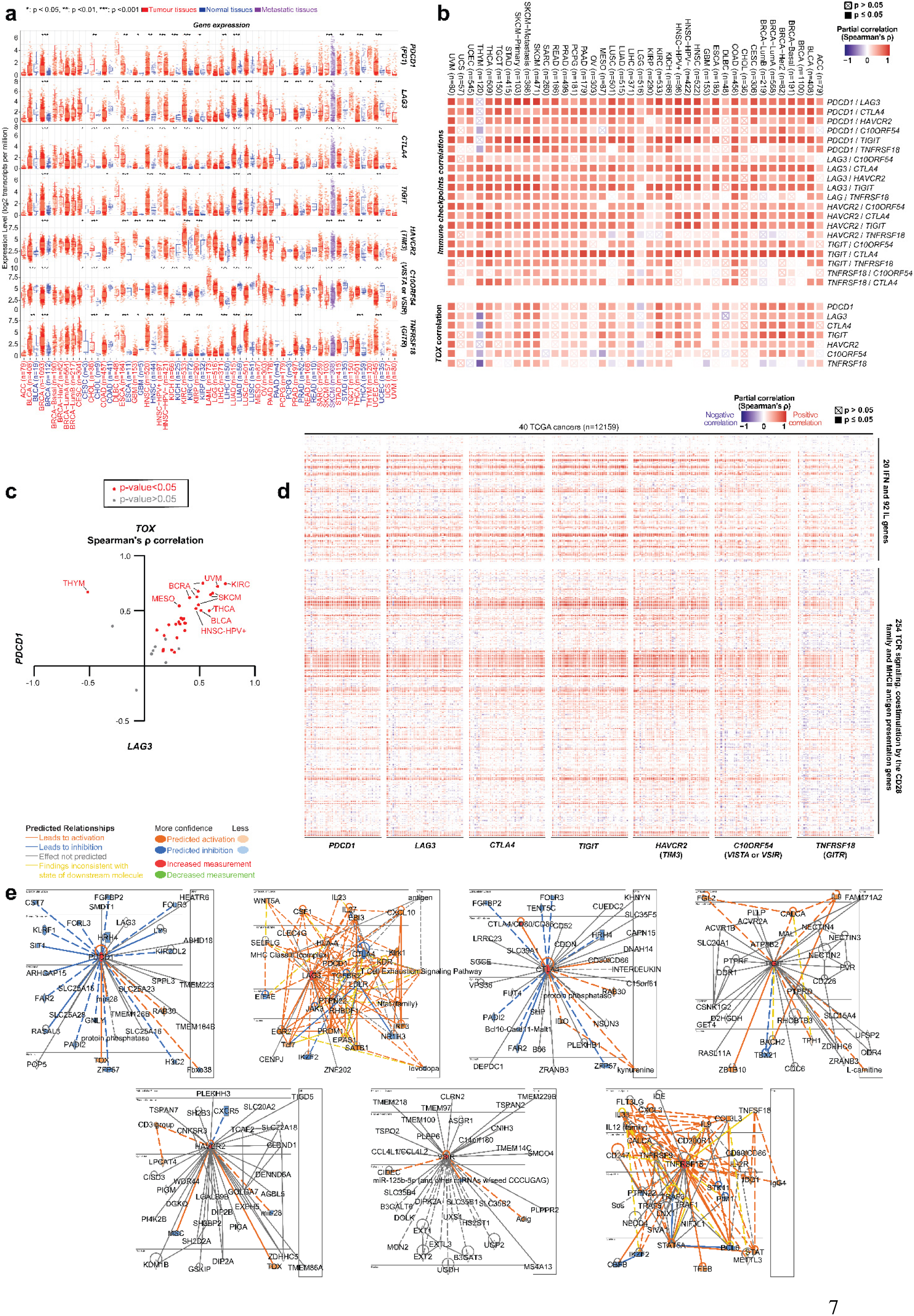
PDCD1, LAG3, TIGIT, TIM3, VISTA and GITR expression, associated gene profiles and regulatory networks in all represented human cancers in the TCGA database. Data of 40 cancer types with a total sample size of n=12159. (a) Median expression of the indicated IC genes among human TCGA cancers. Red, blue, purple indicate tumour, normal and metastatic tissue. Relevant statistical results are shown within the graphs.*, **, ***, indicate significant (p<0.05), very significant (p<0.01) and highly significant differences (p<0.001). (b) Heatmaps represent correlations between IC pairs in each cancer type as indicated, and with TOX gene expression using Spearman’s correlation test as indicated. Crossed squares indicate no significant correlation (p>0.05). (c) Correlation of LAG3 and PDCD1 gene expression with TOX in TCGA cancers as indicated. Red and gray indicate significant (p<0.05) and non-significant (p>0.05) correlations. (d) Heatmap of correlation of the indicated IC gene expression with selected immune genes including IFN, IL, TCR signalling, CD28 co-stimulatory family and MHCII antigen presentation. Detailed information and statistical significance for each specific gene associated to the PDCD1/LAG3 gene signature is shown in supplementary figure S2 and S3 (e) Predicted functional interactome networks after applying IPA algorithms on data from curated publicly available datasets as indicated in the text. Key nodes are shown, and inter-nodal lines represent functional relationships between nodes. In red and orange, up- regulated nodes and interactions. In blue, down-regulated nodes and interactions. Specific legends to inter-nodal relationships are described in IPA (materials and methods).

Independently from analyses of actual genomic data with TIMER2.0, we generated relevant interactome networks associated to the upregulation of each separate IC using IPA. This algorithm reconstructs predicted pathways using input data from datasets of RNA-seq, small RNA-seq, metabolomics, proteomics, microarrays including miRNA and SNP, and small-scale experiments (Kramer et al., 2014). The output networks for each IC were found to regulate similar functions including immune responses, inflammation, cell-to-cell signalling and proliferation **(Figure 1e)**. But notably, *PDCD1*/*LAG3* were the only immune checkpoint pair predicted by IPA to be co-upregulated.

We extended correlations of IC expression with additional 1038 genes involved in cell cycle, cell-cell communication, chromatin organization, DNA repair, DNA replication, programmed cell death and autophagy using global purity-adjusted partial Spearmańs test. Again, we confirmed distinct gene signature profiles associated to particular IC types such as *PDCD1/LAG3/CTLA4*, *TIM3 /TIGIT*, *GITR* and *VISTA* **(Figure 2)** as previously found with only immune-related genes **(Figure 1d)**.

**Figure 2.**
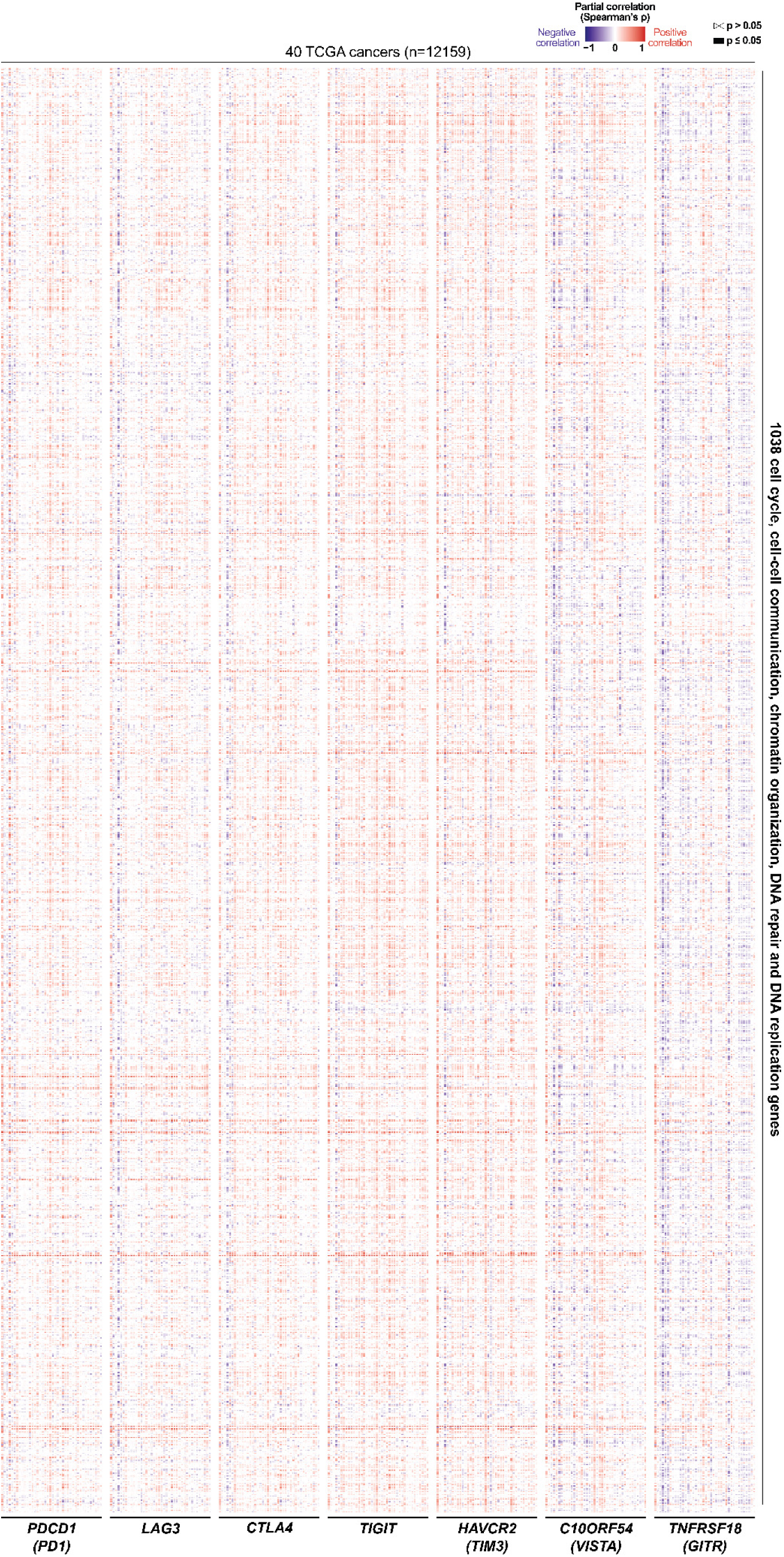
Correlation of PDCD1, LAG3, CTLA4, TIGIT, TIM3, VISTA and GITR IC gene expressions with a selection of genes regulating cell cycle, gene expression and signalling in genomic samples from TCGA cancers. Data from 40 cancer types, total sample size of n=12159. Panels represent heat-map correlates with partial purity-adjusted Spearman’s correlation test for each of the indicated IC gene. Red, positive correlation; Blue, negative correlation. Detailed information and statistical significance for each specific gene associated to the PDCD1/LAG3 gene signature is shown in supplementary figure S5.

### The *PDCD1*/*LAG3*-associated gene signature identified in tumour immune infiltrates corresponds to dysfunctional T-cells

TIMER 2.0 was used to estimate the degree of infiltration with specific immune cell types. Then, correlations with gene expression were calculated with purity-adjusted Spearmańs tests **(Figure S1)**. As expected, and in agreement with our original selection criterion, *PDCD1* and *LAG3* positively correlated with T-cell infiltration including CD4 and CD8 T-cells at several differentiation stages, follicular helper and regulatory CD4 T-cells; and other lymphocyte types such as gamma/delta T-cells, activated natural killer cells (NKs), and naïve, memory and class-switched memory B cells **(<u>Figure</u> <u>S1a</u>)**. In contrast, *PDCD1* and *LAG3* signatures negatively correlated with infiltration of naïve non-regulatory CD4 T-cells, resting NK cells, and common lymphoid progenitors. *PDCD1* and *LAG3* positively correlated with infiltration of myeloid and plasmacytoid dendritic cells (pDCs), M1 and M2 macrophages and monocytes **(<u>Figure S1b</u>)**. Interestingly, a negative correlation was obtained with neutrophils, M0 macrophages, myeloid-derived suppressor cells (MDSCs), mast cells and tumour-associated fibroblasts. These results suggested that tumours with *PDCD1/LAG3* signatures present an inflamed microenvironment, highly infiltrated with non-naïve T-cells and with other pro-inflammatory immune cells and antigen-presenting cells.

*PDCD1* and *LAG3* gene expression was correlated with all gene data available in TCGA cancer tumour immune infiltrates samples as summarized in **(Figure 1, 2)**, encompassing about 369 genes of T-cell function, 114 interleukin and interferon family genes, and other 255 immune related genes. A positive correlation with most interleukin and interferon genes was found, especially with *IL21R*, *IL2RA*, *IL2RB*, *IL2RG*, *IL4I1*, *IL10RA*, *IL15RA*, *IL18BP*, *IL18R1*, *IL12RB1*, *IL12RB2*, *IL18RAP* and *IFNG* **(Figure 1d<u>, S2, S3</u>)**. As expected for T-cell infiltrates, the *PDCD1/LAG3* signature correlated with regulators of TCR signalling (*CD4, CD3E, CD3D, CD3G, CTLA4, ICOS, ZAP70, LAT, LCK*), CD28 family costimulation molecules (*CD28, CD80, CD86, CARD11, CD101, CD247, CD74*) and MHC II antigen presentation (*CTSS, FYB, BYN, GRAP2),* multiple *HLA* genes, among others **(Figure S3)**. Overall, these results demonstrated the association of *PDCD1/LAG3* signatures with inflamed, immune-infiltrated tumours. An exception to the rule was thymoma that showed the opposite correlation for *PDCD1* and *LAG3* expression with almost all the studied genes.

To identify the functional pathways associated to the immune genes correlating with *PDCD1* and *LAG3* gene signatures, this omic co-signature was reconstructed as regulatory protein networks and causal relationships with IPA and compared to signatures associated to each IC separately (Kramer et al., 2014) **(Figure 3)**. The PD-1/LAG-3 signature in human cancers corresponded to T-cell exhaustion pathways and PD-1/PD-L1 blockade immunotherapy **(Figure 3a)**. This signature was also associated to inositol metabolism and super-pathway of inositol phosphate compounds. Specific signalling and antigen-presentation molecules were identified as causal relationships **(Figure 3b)**. The most significant were PTPN22, KDR, EBI3, SLPLG, NFAT family, CD80, CD274, ADORA2A, GSK3A and IL-1R. A cascade of upstream transcriptional regulators was found associated to the PD-1/LAG-3 signature **(Figure 3c)**. Some of these are well-known T-cell inhibitors associated to immune checkpoints and PD-1 signalling such as PTPN22, CTLA-4, VEGFA, SATB1 and FOXP3 among others. The PD-1/LAG-3 co-signature was linked to bio-functions, toxicities and diseases characteristic of immune dysfunctionality and cancer **(Figure 3d, 3e)**. These results confirmed that PD-1/LAG-3 signatures in human cancers represented gene expression profiles associated to T-cell dysfunctionality.

**Figure 3.**
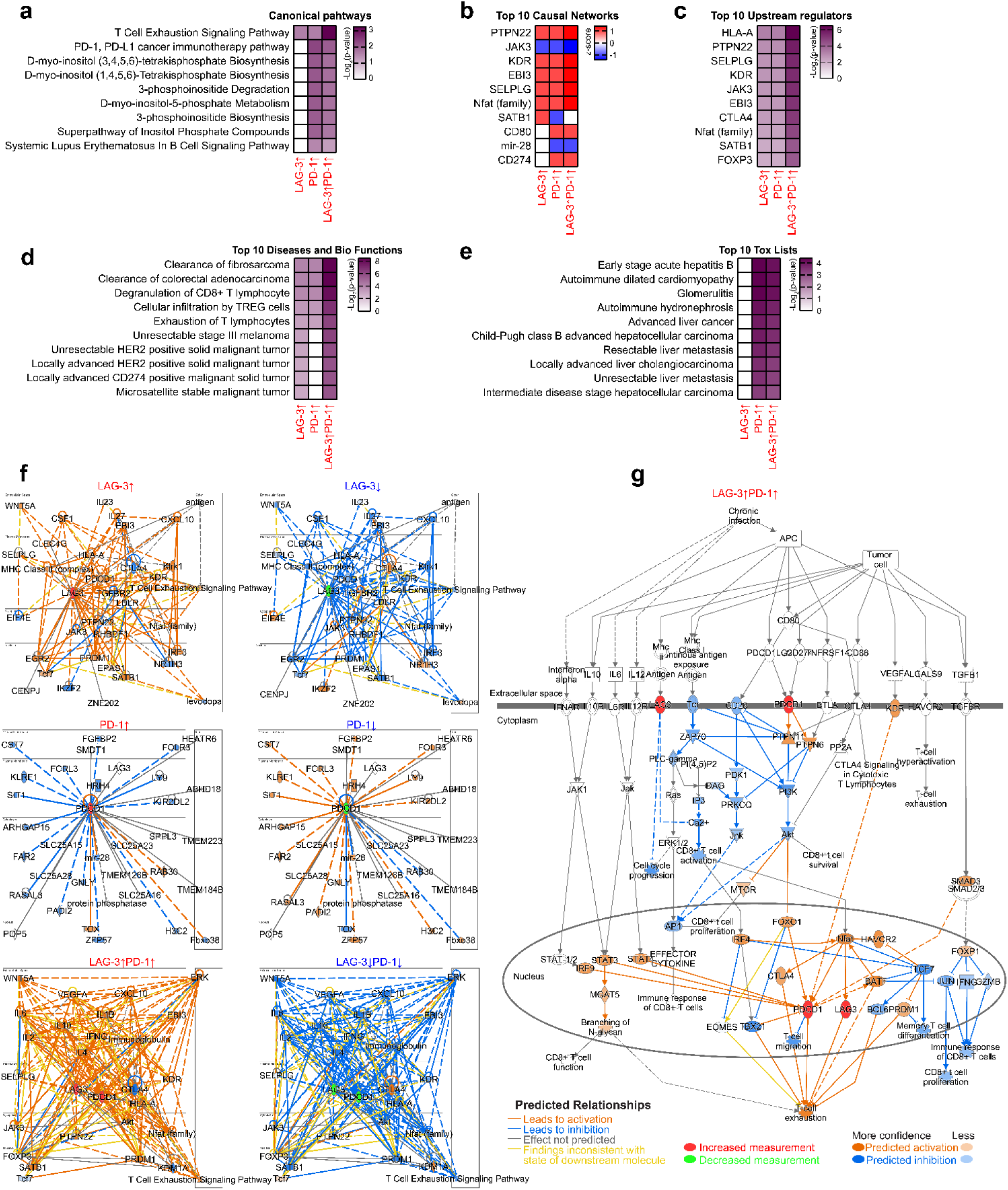
Regulatory networks and causal relationships associated with PD-1 / LAG-3 signature. QIAGEN IPA algorithms were applied on data from curated publicly available datasets of RNA-seq, small RNA-seq, metabolomics, proteomics, microarrays including miRNA and SNP, and small-scale experiments. **(a)** The intensities in enrichment of the indicated canonical pathways represented as up- regulation of PD-1/LAG-3 and combinations **(b)** Top 10 identified causal networks as indicated are correlated with z-scores to the up- or down-regulation of PD-1/LAG-3 and combinations as shown in the figure (red, up-regulated; blue, down-regulated). **(c)** Intensities of enrichment of the indicated top 10 upstream regulators represented as a function of the up- or down-regulation of PD-1/LAG-3 and combinations. **(d)** Intensities of enrichment of the indicated top 10 diseases and biofunctions as a function of the up- or down-regulation of PD-1/LAG-3 and combinations. **(e)** As in (d) but with the top 10 tox lists. **(f)** Predicted regulatory interactomes and associated networks with the indicated PD-1 and LAG-3 signatures (up-regulated in red, down-regulated in blue). Key nodes are shown, and inter- nodal lines represent functional relationships between nodes. In red and orange, up-regulated nodes and interactions. In blue, down-regulated nodes and interactions. **(g)** Reconstructed regulatory interactomes and associated networks for the T-cell exhaustion pathway signalling and TCR downregulation associated to both PD-1 and LAG-3 upregulation. The specific legends to inter-nodal relationships are described in IPA (<u>Ingenuity Pathway Analysis | QIAGEN Digital Insights</u>).

We also modelled individual interactome networks for activated or inhibited PD-1 and LAG-3 pathways using IPA, and compared them to the omics co-signature as found in human cancers **(Figure 3f)**. PD-1 or LAG-3 individually showed signalling networks that clearly differed when compared to the PD-1/LAG-3 co-signature, evidencing the relevance of co-signalling in accentuated T-cell dysfunctionality **(Figure 3f)**. Again *in-silico* modelled signalling networks associated to the PD- 1*/*LAG-3 signature were identified as T-cell exhaustion. We previously described in mouse T-cells that CD3 down-modulation is characteristic of PD-1 signalling after PD-L1 engagement in T-cells (Karwacz et al., 2011). This was confirmed in the modelled pathways from the omics data, which highlighted networks leading to downregulation of TCR signalling and termination of signal transduction **(Figure 3g)**.

IPA-reconstructed pathways were supported by omics data associated to *PDCD1* and *LAG3* gene signatures in TCGA cancers, as shown by correlation of this signature with most regulators predicted by IPA **(Figure S4)**. These correlations were particularly strong for *CTLA4, CSF1, CXCL10, EBI3, IL27, KLRK1, PRDM1, PTPN22, SEPLG, HLA-genes, FOXP3, IFNG, IL10, ARHGAP15, CST7, FCRL3, GNLY, LY9, RASSAL3, SIT1, SLC24A28* and *TOX*.

**Figure 4.**
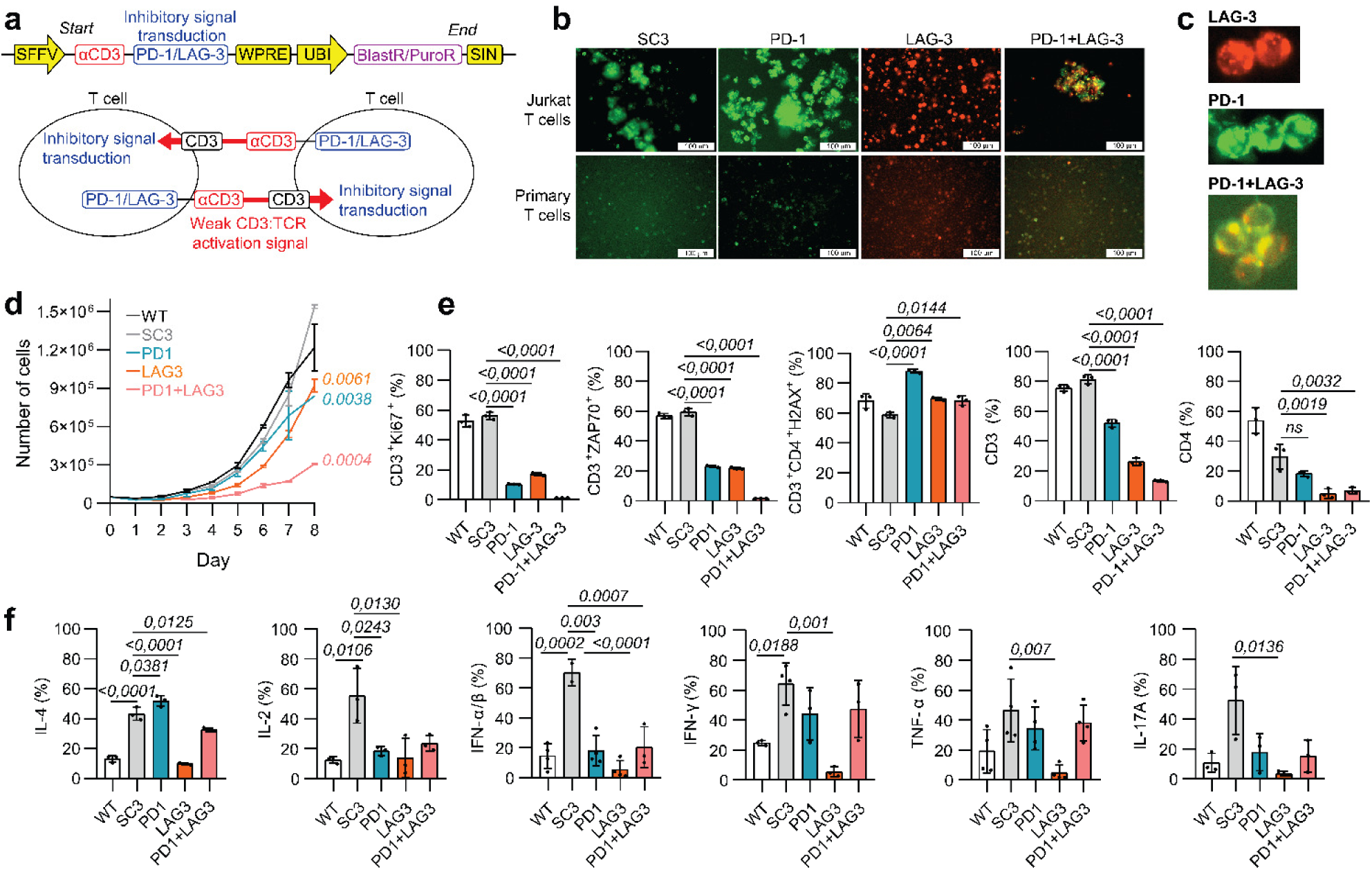
Expression of molecules with constitutive PD-1 and LAG-3 signalling in human T-cells. (a) Top, pDUAL lentivector expressing the fusion genes (SC3-PD-1 or SC3-LAG-3) and blasticidin or puromycin resistance, as indicated. The anti-CD3 single-chain antibody (SC3) coding sequence fused to LAG-3 or PD-1 stem-transmembrane-intracellular regions is schematically shown. Bottom, mode of action of the engineered molecules expressed by two neighbouring T-cells. The SC-3 domain binds and delivers a weak signal to the corresponding TCR through CD3 binding. PD-1 and LAG-3 signalling domains deliver the corresponding inhibitory signals. SFFV, spleen focus-forming virus promoter; LTR, long-terminal repeat: UBI, human ubiquitin promoter; SIN, self-inactivating LTR; WPRE, woodchuck post-transcriptional response element; PuroR, puromycin resistance gene; BlastR, blasticidin resistance gene. **(b)** Fluorescence microscopy pictures of living Jurkat and primary T-cells expressing SC3-PD-1-GFP (green), SC3-LAG-3-Cherry (red) or both (yellow). **(c)** Detail of engaging T-cells expressing the constructs. **(d)** Growth of Jurkat T-cell lines expressing the indicated constructs (n=3; error bars, standard deviations). Relevant statistical comparisons between the cell lines and control cell lines are shown. **(e)** Bar graphs of T-cell percentages expressing CD3, CD4, Ki67, ZAP70 and a DNA damage marker (pS139-H2AX) within the indicated T-cell lines (n=3; error bars, standard deviations). **(f)** Bar graphs of T-cell percentages expressing the indicated cytokines within the T-cell lines shown in the graphs (n=4; error bars, standard deviations). Means are represented together with standard deviations as error bars. Two-way ANOVAs were carried out, followed by a posteriori pair- wise comparisons with Tukey’s test.

The *PDCD1/LAG3* gene expression profile was further correlated with additional 1038 non-immune-related genes, including regulators of cell cycle, cell-cell communication, chromatin organization, DNA repair, DNA replication, programmed cell death and autophagy **(Figure 2<u>, S5</u>)**. Overall, strong correlation with cancer-altered gene regulation profiles was found for most of the studied genes **(Figure S5).** Again, an association of these regulators was found with downstream targets which negatively regulated the TCR signalosome, including genes involved in immune synapse formation, TCR-associated kinases, phosphatases, cytokine signalling kinases and PI3K/AKT signalling pathway, among others **(Figure S5)**. Remarkably, *PDCD1/LAG3* showed similar correlation patterns for almost of all the studied molecules (>1000) except for thymoma **(Figure 1, 2<u>, S1-S5</u>)**. THYM samples showed negative opposite correlations for *PDCD1* and *LAG3* expression for almost all the studied molecules, clearly evidencing a distinct mechanism of response in this tumour towards PD-1/LAG-3 co-signalling **(Figure 2<u>, S2-S5</u>)**.

### Establishment of human T-cell lines with constitutive PD-1 and LAG-3 signalling

Our results of *PDCD1/LAG3* omics signature in human cancers uncovered gene expression profiles and functional interactome networks associated to cancer cell markers and to T-cell dysfunctionality. To compare these signatures with those in T-cells with activated PD-1/LAG-3 signalling pathways, we induced PD-1/LAG-3 signalling either separately or in combination to compare their profile to that from our omics studies. To that end, PD-1 and LAG-3 molecules with constitutive signalling activities in T-cells were engineered. PD-1 and LAG-3 extracellular immunoglobulin-like domains were replaced with a CD3-binding single chain antibody (SC3) previously described by us (Zuazo et al., 2019). When expressed in T-cells, the SC3 domain binds to CD3 from neighbouring T-cells. This interaction simultaneously triggers TCR signal transduction and signalling through the intracellular domains of PD-1, LAG-3 or both (PD-1+LAG-3 cell lines) **(Figure 4a)**. These fusion genes (termed SC3-PD-1 and SC3-LAG-3) were cloned into lentivectors for T-cell transduction. As a control, the SC3 molecule without PD-1 or LAG-3 intracellular tails was used (Zuazo et al., 2019). To first test their expression in T-cells, GFP and mCherry were fused to their carboxy-termini. Both PD-1 and LAG-3-based constructs showed good expression levels and accumulated at T-cell-to-cell contacts in human primary cells and Jurkat CD4 T-cells **(Figure 4b, 4c)**. However, it was not possible to maintain primary T-cells in culture alive when expressing these constructs. The growth of Jurkat T-cells expressing either of the constructs was noticeably delayed compared to control cell lines, especially for the PD-1+LAG-3 co-expression **(Figure 4d)**. The same constructs without GFP or mCherry exerted the same inhibitory effects and were chosen for further studies to eliminate artefacts caused by the fluorescent proteins.

To evaluate whether constitutive PD-1 and LAG-3 signalling singly or in combination (PD-1+LAG-3) caused differential T-cell phenotypes, 30 cell markers were analysed representing indicators of T-cell dysfunctionality, differentiation, proliferation, and DNA damage **(Figure 4e, 4f, Figure S6)**. PD-1, LAG-3 and PD-1+LAG-3 showed distinct differentiation phenotypes related to inhibitory functions. An increase in central memory (CM) populations was observed through PD-1 or LAG-3 signalling **(<u>Figure S6a</u>)**, with the exception of PD-1/LAG-3 combination which showed increased effector memory (EM) phenotypes. A strong inhibition of proliferation was again evidenced by reduced Ki67 expression, and key T-cell markers such as CD3 and CD4 were downregulated **(Figure 4e)**. ZAP70 expression was evaluated as a marker of TCR signalling capacities, which was strongly reduced by PD-1 or LAG-3 signalling, and nearly completely abrogated by the PD-1+LAG-3 combination **(Figure 4e)**. DNA damage was increased specially with PD-1 signalling in T-cell cultures, as assessed by pS139-H2AX detection **(Figure 4e)**. CD69 and endogenous PD-1 were upregulated with PD-1 signalling **(<u>Figure S6b</u>)**. Additionally, expression of all tested Th1, Th2 and Th17 cytokines was downmodulated in all T-cell lines expressing PD-1, LAG-3 or PD-1/LAG-3 with some exceptions **(Figure 4f<u>, Figure S6c</u>)**.

### Functional interactomes caused by constitutive PD-1, LAG-3 and PD-1+LAG-3 signalling regulate molecular dysfunctionality in T-cells

Proteomes of all engineered T-cell lines were analysed by quantitative differential proteomics. Three independent cultures (biological replicates) were used for each cell line with the exception of PD-1 and PD-1+LAG-3 (two independent cultures). A total number of 2854 proteins was identified of which 833 (p≤0.05) and 539 (p≤0.01) were differentially expressed **(Figure 5a, 5b)**. The top enriched canonical pathways and biological processes for the differential proteomic dataset were related to RNA metabolism and cell signalling pathways including EIF2, FAT10, BAG2 and mTOR pathway, inhibition of ARE-mediated mRNA degradation pathway, mitochondrial dysfunction, protein ubiquitination pathway, sirtuin signalling pathway, and regulation of eIF4 and p70S6K signalling **(Figure 5c<u>, S7a, S7b</u>)**. Functional interactomes corresponded to altered downstream TCR signal transduction pathways, including NF-kB signalling, cell growth through mitotic cell cycle checkpoints and cytokine signalling. The identified top 5 upstream regulators of the identified proteomic dataset were RICTOR, MYC, LARGP1, TP53 and HNF4A **(Figure 5d)**. The top 5 causal networks of the identified proteomic dataset were TFDP1, MKNK1, CDC20, TOP2 and MTORC2. These upstream regulators and causal network molecules regulate cell growth, proliferation, gene expression, protein synthesis and response to environmental stress and cytokines. Reactome, Metascape, STRING, and IPA analyses confirmed these observations **(Figure 5<u>, S7-S12</u>)**.

**Figure 5.**
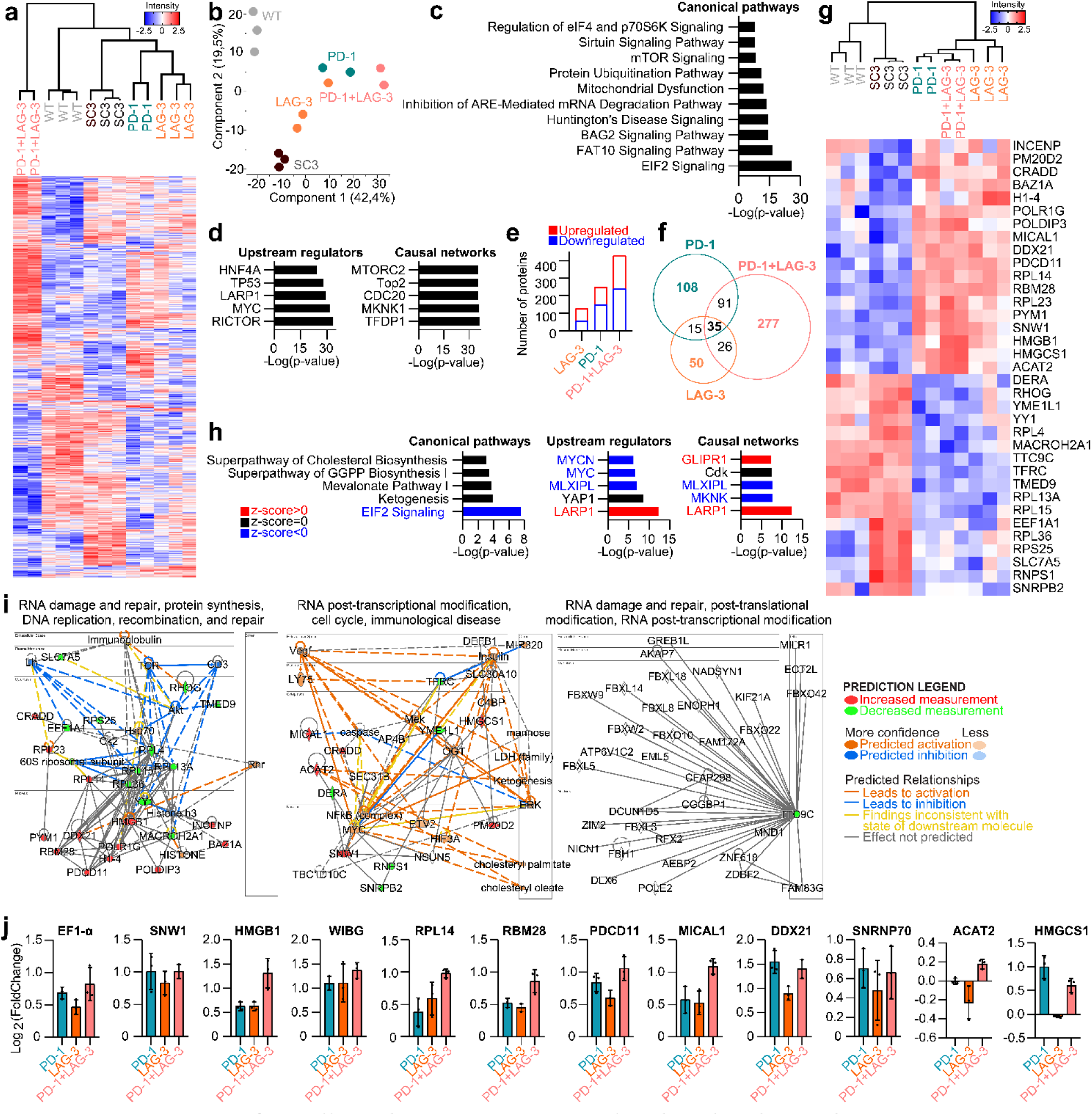
Proteomes of T-cells with PD-1/LAG-3-regulated molecular pathways. (a) Heat map of regulated proteins from Jurkat T-cell lines with active PD-1, LAG-3 pathways, or their combination, together with unmodified and SC3 cell lines as controls. Red, upregulated proteins; blue, down- regulated proteins. **(b)** Principal component analysis of the indicated individual samples. **(c)** Top 10 canonical pathways associated with the differential proteomic profiles. **(d)** Top 5 upstream regulators and causal networks associated with the differential proteomic profiles. **(e)** Column graph with the number of differentially regulated proteins in PD-1, LAG-3, and PD-1+LAG-3 Jurkat T-cell lines compared to SC3 control cells (p-value ≤ 0.05) for up-regulated (Red, Log2 Fold Change) ≥ 0.38) and downregulated (Blue, Log2 Fold Change ≤ -0.38) proteins. **(f)** Venn diagram of differentially regulated proteins in the indicated T-cell lines compared to SC3 control cells as a common standard. 35 common proteins were regulated for all conditions. **(g)** Heat map of expression of the 35 common proteins as in (f) within the proteomes of all T-cell lines. **(h)** Column graphs for the top 5 canonical pathways (left), upstream regulators (centre) and causal networks (right) associated with the 35 commonly regulated proteins. **(i)** Three top functional interactome networks (functions indicated on top of each network) associated to the 35 commonly regulated proteins. The specific legends to inter-nodal relationships are described in IPA (**<u>Ingenuity Pathway Analysis | QIAGEN Digital Insights</u>**).**(j)** Bar graphs of transcriptional transactivation of the indicated gene promoters identified as regulated by PD-1/LAG-3 signalling in T-cell lines. Promoters expressing GFP were introduced by lentivector transduction in T-cell lines expressing constitutively active PD-1, LAG-3 or PD-1+LAG-3 molecules as indicated. Transactivation was quantified by GFP expression. Means from 3 independent repetitions are shown, together with standard deviations as error bars.

To identify functional interactomes specifically triggered by PD-1, LAG-3 and PD-1+LAG-3 signalling, we used the proteome of SC3 control cells as a common comparative standard **(Figure 5e, 5f<u>, S7a, S7e-g</u>)**. The ontology terms were common for all the cell lines, but PD-1+LAG-3 T-cells showed the most divergent proteomes with the highest number of differential proteins **(Figure 5a, 5e-f<u>, S7a</u>)**. Indeed, a major part of the regulated proteome was unique for this combination, which was an indication of a PD-1/LAG-3-regulated specific programme **(<u>Figure S7a, S7e-c</u>)**. RNA processing, metabolism and intracellular transport were specially altered by PD-1+LAG-3 co-signalling **(<u>Figure</u> <u>S7g, S8-S12</u>)**. Only 35 proteins were found to be significantly commonly regulated for PD-1, LAG-3, and PD-1+LAG-3 proteomes (17 were upregulated and 18 downregulated) **(Figure 5f-g)**. These three PD-1, LAG-3, and PD-1/LAG-3 proteomic signatures had LARP1, MLXIPL, MYC and MYCN as common upstream regulators and LARP1, GLIPR1, MLNK and MLXIPL as common causal networks **(Figure 5h**.). In agreement with our computational omics analyses of TCGA cancers, upregulated proteins were involved in RNA processing, epigenetic regulation, histone modifications, apoptosis, and transcription **(Figure 5g-i)**. Interestingly, the commonly regulated 35 proteins were indicators of a significant downregulation of the EIF2 signalling pathway **(Figure 5g-h)**, which in turns inhibits RNA and protein metabolism and TCR signalling **(Figure 5i)**. To test whether some of these targets were transcriptionally regulated, T-cell lines expressing the activated immune checkpoints were transduced with lentivectors expressing GFP under promoters corresponding to a selection of the up- regulated targets in PD-1/LAG-3 proteomes. These targets were involved in RNA and gene regulation **(Figure 5i)**. Increased transcriptional activity was observed for these promoters compared to control cells as evaluated by fold change in GFP expression **(Figure 5j)**. MTA1, RBBP4 and DDXD21 proteomic data was validated by flow cytometry. TFactS, an in-silico prediction algorithm of regulated transcription factors also identified MYC, SREBF1, ATF1, ARNT and CREBBP as the top 5 regulators for the 35 common proteins between cell lines expressing inhibitors **(<u>Figure S7c-d</u>)**.

Then, well-known T-cell characteristics regulated by PD-1 and LAG-3 signalling were studied within our data **(Figure 6)**. An analysis of up and downregulated proteins with PD-1, LAG-3 and PD-1+LAG- 3 activated pathways was compared to the SC3 control cell line, showing similar molecular functions and protein interaction networks. These included protein translation and elongation, rRNA processing and altered splicing pathways **(<u>Figure S8-S11</u>)**. Downregulation of the EIF2 signalling pathway was again the only common canonical pathway for all PD-1, LAG-3 and PD-1+LAG-3 proteomes, followed by NGF, PAK, CXCR3, GTPases, reelin and RAC signalling on PD-1 and PD-1+LAG-3 proteomes **(Figure 6a-c, <u>S7h</u>)**.We previously demonstrated that PD-1 signalling in mouse T-cells caused CD3 downmodulation (Karwacz et al., 2011). Our phenotypic data further extended this data to PD-1, LAG- 3 and PD-1+LAG-3 activation in human T-cells **(Figure 4e)**. Our proteomic data confirmed these experimental findings linking them with a reduction of other TCR signalling regulators **(Figure 5, 6d-e, <u>S8-S12</u>)**, and inhibition of pathways associated to T-cell activation such as TCR, Lh, SYVN1, NFKB, CEBPB and CCDN **(Figure 6d-e)**. Impaired cell growth was also confirmed by proteomic quantification of cell division regulators **(<u>Figure S8c</u>)**. This included upregulation of RB1 expression and CDK1/CDK6 down-regulation in PD-1+LAG-3 co-expressing cells **(<u>Figure S8-S12</u>)**. Importantly, PD-1+LAG-3 co-signalling showed proteomic profiles associated to pronounced cell growth inhibition, which agreed with experimental cell growth rates **(Figure 4d, 4e, 6, <u>S8-S12</u>)**. We found GABA, CDKN2A and LARP1 within the top ten upstream activated regulators in PD-1 and LAG-3 proteomes **(Figure 6b)**. Most of them were also significant causal networks for all proteomes **(Figure 6d)**. As expected, the top tox functions, lists and bio functions associated with the PD-1, LAG-3 and PD-1/LAG-3 proteomes were related to cancer, impaired cell function and immune diseases as shown in **Figure 6f-h**.

**Figure 6.**
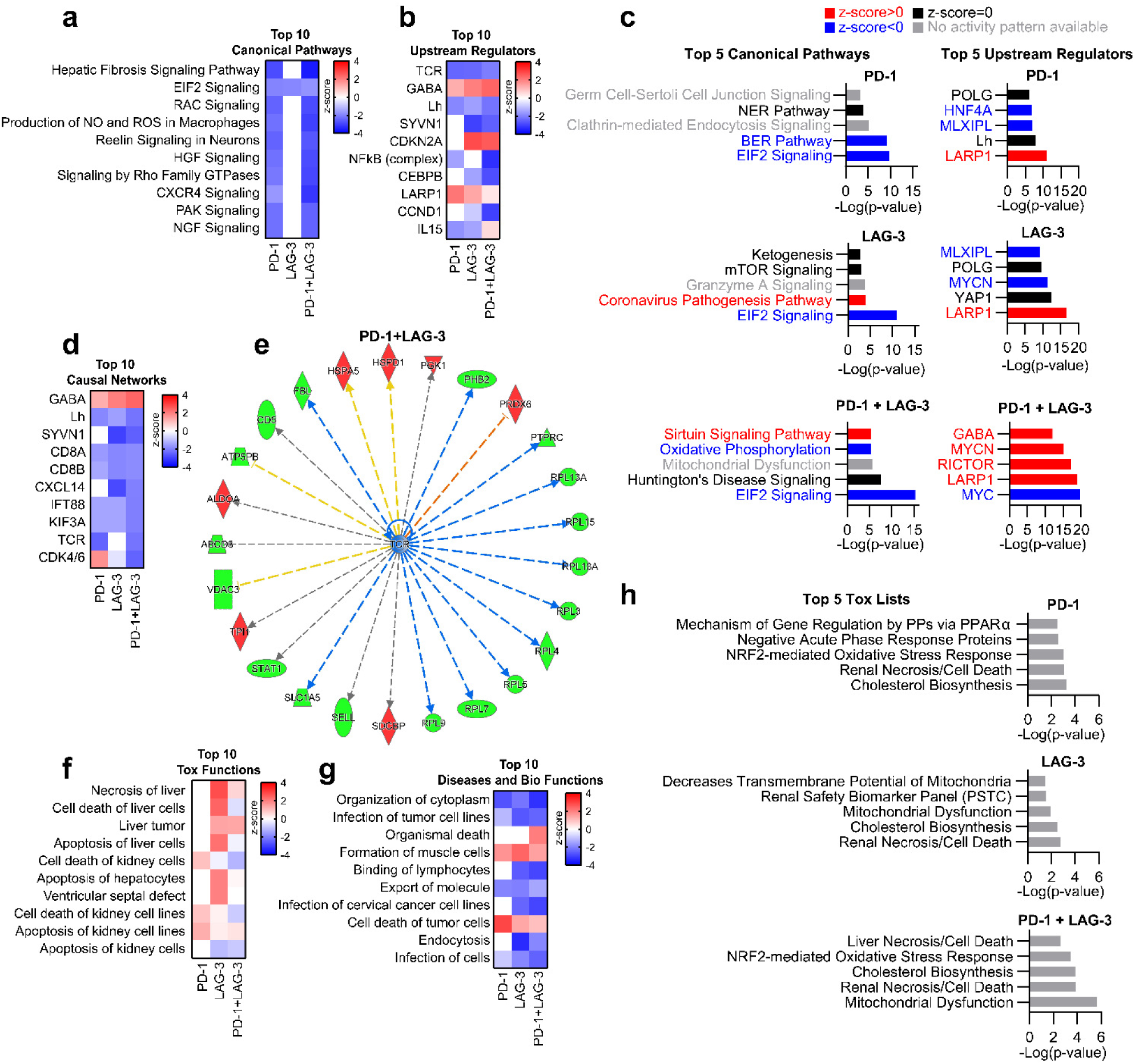
Molecular dysfunctionality programme in T-cells by PD-1/LAG-3 co-signalling. (a) Common top 10 canonical pathways associated with PD-1, LAG-3, or PD-1+LAG-3 proteomes. **(b)** Common top 10 upstream regulators. **(c)** Top 5 canonical pathways and upstream regulators. **(d)** Common top 5 causal networks. **(e)** Regulation of key functional targets linked to TCR downregulation within the PD-1+LAG-3 proteome. IPA analysis identified TCR down-modulation as an upstream regulator of PD-1, LAG-3 and PD-1+LAG-3 T-cell proteomes. The graph shows relationships between the predicted downmodulated TCR (in the centre in blue) and differential expression of the identified proteins in the PD-1+LAG-3 proteome. In red, upregulated proteins. In green, downregulated proteins. Blue lines, predicted downregulation; orange lines, predicted upregulation; grey indicates a predicted relationship with a non-predicted effect, and yellow lines, predicted relationship findings inconsistent with the state of the downstream molecule. **(f)** Common top 10 tox functions. **(g)** Common top 10 diseases and bio functions associated to the indicated proteomes. **(h)** Top 5 tox lists associated. Red, predicted upregulation; blue, predicted downregulation; grey, predicted relationship with a non-predicted effect. The specific legends to inter-nodal relationships are described inIPA (**<u>Ingenuity</u> <u>Pathway Analysis | QIAGEN Digital Insights</u>**).

### Functional overlaps of PD-1/LAG-3 omic signatures in human cancers with PD-1+LAG-3 T-cell proteomes uncover common dysfunctionality pathways

The proteomes uniquely associated to PD-1+LAG-3 Jurkat T-cells corresponded to the regulatory pathways in *PDCD1/LAG3* gene signatures found in TCGA human cancers which were associated to T-cell functions **(Figure S13)**. The top enriched pathways were linked to down-regulation of RNA metabolism, particularly nonsense-mediated decay (NMD), rRNA processing, and down-regulation of protein metabolism through alterations of eukaryotic translation regulators and membrane targeting. As expected from our previous analyses, pathways associated to TCR signal transduction were strongly down-modulated, exemplified by CD3epsilon downmodulation and functional networks leading to LCK inactivation and PD-1-dependent dephosphorylation of TCR chains, as detailed in **<u>Figures S8-</u> <u>S12</u>**. Other processes previously identified were also confirmed, such as regulation of various cell cycle checkpoints, modulators of G1/S DNA damage, G2/M transition and formation of mitotic spindles **(<u>Figures S8-S12</u>)**. Reduced expression of CDK1 and CDK6 was evident, together with pathways leading to Cdh1 degradation. PD-1/LAG-3 signatures in T-cells were consistent with enriched chromosome maintenance pathways by decreased expression of H2 clustered histones. Some of these pathways included the non-extension of telomeres, and suppression of epigenetic regulation by differential expression of transcriptional and chromatin regulators **(<u>Figures S8-S12</u>)**. In addition, PD- 1+LAG-3 specific proteomes in T-cells and TCGA cancers were consistent with upregulation of extrinsic and intrinsic apoptotic pathways, their execution phases, but also activation of autophagy as previously shown for PD-1 signalling (Arasanz et al., 2017; Wang et al., 2019; Wu et al., 2020; Wu et al., 2021). Importantly, the downregulation of the EIF2 signalling pathway was found to be commonly regulated in T-cell proteomes and omics data from TCGA cancers, as evidenced by downmodulated EIF2B2, RPL4, RPL5 or RPL7 protein expression (**Figure 7a**). Overall, these results confirmed experimental findings by us and others. But importantly, our data uncovered novel pathways of epigenetic/transcriptional regulation of T-cell dysfunctionality associated to the PD-1/LAG-3 co-signalling signature both in T-cell lines and in omic TCGA data.

**Figure 7.**
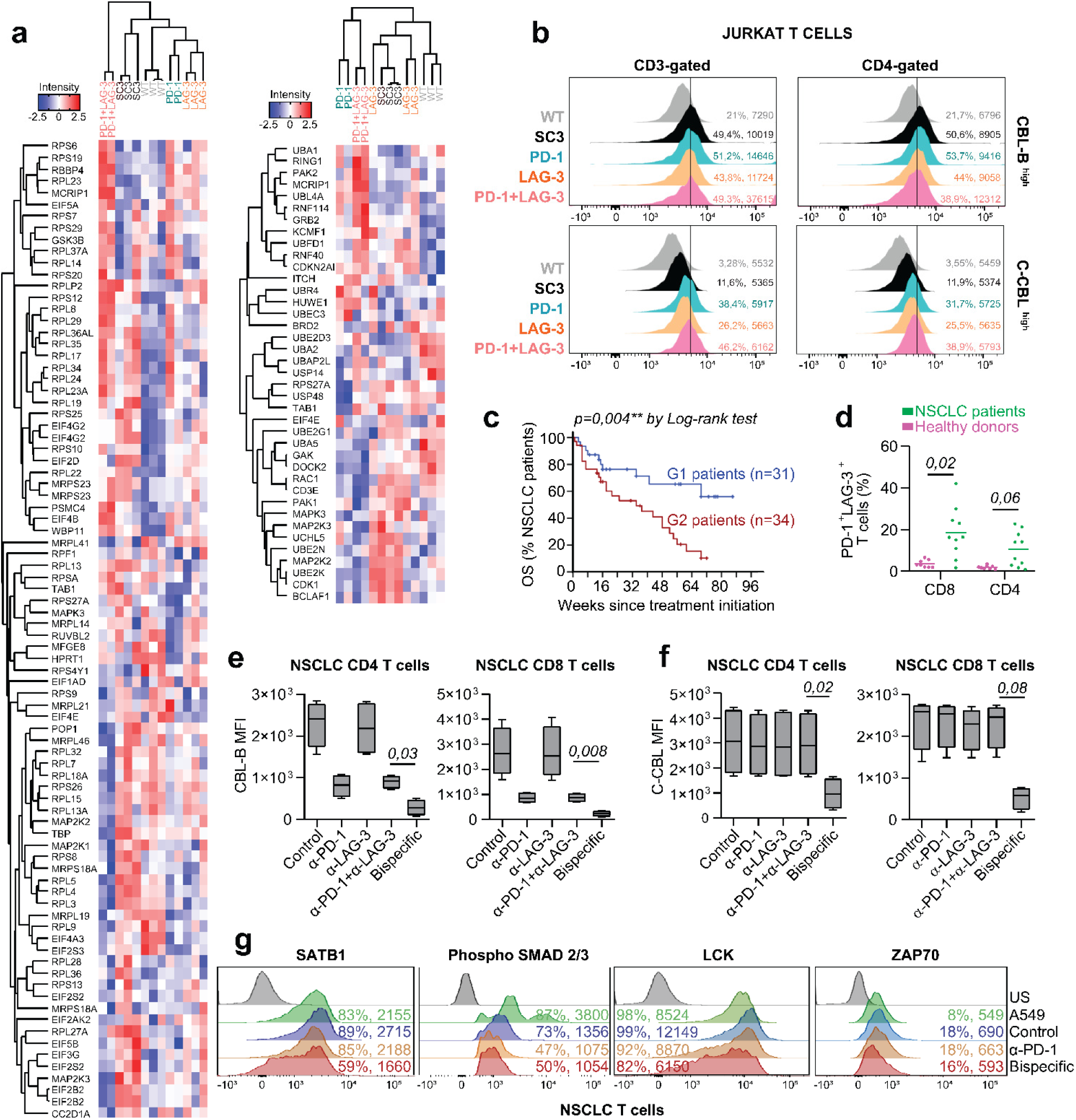
CBL E3 ubiquitin ligases as key targets for PD-1/LAG-3 co-blockade. (a) Heat map of statistically significant differential proteins of the EIF2 signalling pathway (left), and E3 ubiquitin signalling proteins (right) within the indicated T-cell proteomes. Red, significantly up-regulated proteins; Blue, significantly down-regulated proteins. **(b).** Flow cytometry histograms of CBL-B and C-CBL expression in the indicated T-cell lines. Percentages of positive cells and mean fluorescent intensities (MFIs) are plotted. **(c)** Overall survival of NSCLC patients treated with PD-L1/PD-1 blockade monotherapies, separated into G1 and G2 groups. G1 patients present CD4 T-cells without co-upregulation of PD-1/LAG-3, while G2 patients present dysfunctional PD-1+/LAG-3+ CD4 T-cells. **(d)** Dot plot with the percentage of CD4 and CD8 T-cells that co-express PD-1 and LAG-3 after ex vivo activation, from healthy donors (n=8) and NSCLC patients (n=10). Statistical comparisons were performed by the Mann-Whitney test. Statistical significance was tested by Log-Rank test. **(e)** CBL-B expression by mean fluorescent intensities in CD4 and CD8 T-cells from a sample of non- responder NSCLC patients (n=4), activated ex vivo in the presence of the indicated treatments. Statistical comparisons were carried out by a two-way ANOVA to eliminate inter-patient variability followed by pair-wise Tukey tests. **(f)** Same as **(e)** but for C-CBL expression. **(g)** Flow cytometry histograms of SATB1, Phospho SMAD 2/3, LCK and ZAP70 expression. Gates were established according to unstained controls in T-cells from a sample of non-responder NSCLC patients (n=4).

### CBL E3 ubiquitin ligases are key inhibitory regulators targetable by PD-1/LAG-3 bispecific co- blockade in primary T-cells from lung cancer patients

Our data identified CBL ubiquitin ligases as regulators of a common pathway controlling EIF2 and downregulation of TCR signaling pathways by PD-1/LAG-3 co-signalling (Figure 5,6). CBL-B and C-CBL are the main members of this family of potent inhibitors of T-cell functions (Bachmaier et al., 2000; Chiang et al., 2000; Karwacz et al., 2011; Naramura et al., 2002; Nurieva et al., 2006; Shamim et al., 2007). These features were displayed in our T-cell lines both by proteomics and flow cytometry **(Figure 7a, b)**. CBL proteins and downstream pathway molecules were similarly regulated in human cancers with a *PDCD1/LAG3* signature in our omics data **(<u>Figure S14a</u>)**. To reconstruct CBL-B and C-CBL signalling networks, IPA algorithms were applied **(Figure S14)**. CBL proteins, EIF2 pathway and downstream pathway molecules were differentially regulated in human cancers and in T-cells always in association with *PDCD1/LAG3* gene and PD-1/LAG-3 proteomic signatures **(Figure 7a-b**, **<u>Figure S14a</u>)**. CBL-B and C-CBL expression were identified with immunotherapy and T-cell exhaustion canonical pathways **(<u>Figure S14b</u>)**. T-cell functions were affected at multiple levels **(<u>Figure S14b-c</u>)**. Among these, cell cycle progression was inhibited by C-CBL activation **(<u>Figure</u> <u>S14d</u>)**.

To find out whether CBL proteins are a target for PD-1 and LAG-3 co-blockade, T-cells from NSCLC patients resistant to PD-L1/PD-1 blockade immunotherapies were isolated. These patients showed drastically reduced overall survival **(Figure 7c)** and their T-cells strongly co-upregulated PD-1 and LAG-3 following *in vitro* stimulation with human lung cancer A549-SC3 stimulator cells **(Figure 7d)** as described in (Edwards et al., 2021; Zuazo et al., 2019). The capacities of PD-1/LAG-3 blockers to restimulate these dysfunctional T-cells were assessed by co-culturing them with A549-SC3 stimulator cells. Co-cultures were carried out in the presence of an isotype control, anti-PD-1, anti-LAG-3 and anti-PD-1+anti-LAG-3 antibodies. A bispecific molecule in clinical development (Humabody CB213) was incorporated in the study. This bispecific molecule simultaneously blocks PD-1 and LAG-3 in molecular proximity (Chocarro et al., 2022a; Edwards et al., 2021). PD-1 blockade inhibited CBL-B up-regulation in T-cells from NSCLC patients **(Figure 7e)**, confirming results in mouse T-cells (Karwacz et al., 2011). In contrast, LAG-3 blockade alone did not interfere with CBL-B upregulation. Co-blockade with anti-PD-1 and anti-LAG-3 antibodies inhibited CBL-B expression similarly to PD- 1 blockade alone. But interestingly, co-blockade with CB213 was the most potent inhibitor of CBL-B expression **(Figure 7e)**.

Previous experimental data showed that PD-1 blockade did not inhibit C-CBL expression (Karwacz et al., 2011). In agreement with this, LAG-3 blockade alone, or the anti-PD-1 plus anti-LAG-3 combination did not block C-CBL expression in CD4 and CD8 T-cells from NSCLC patients. In contrast, CB213 significantly blocked C-CBL expression **(Figure 7f)**. SATB1, and LCK were also downregulated as evaluated by flow cytometry following co-blockade with CB213 **(Figure 7g)**. Importantly, SATB1 and SMAD2/3 downregulation after PD-1/LAG-3 bispecific blockade was consistent with the modelling of PD-1/LAG-3 co-expression signature **(Figure 3g)**. Overall, these results suggested that PD-1 and LAG-3 co-blockade by a single bispecific molecule achieves both CBL-B and C-CBL down-regulation in T-cells, unlike the combined use of separate blocking monoclonal antibodies. These results supported that PD-1 and LAG-3 co-signalling regulates a specific molecular programme that enhances dysfunctionality in T-cells in cancer.

## DISCUSSION

Co-upregulation of LAG-3 with PD-1 is a major mechanism of resistance to conventional PD-L1/PD- 1 blockade (Grosso et al., 2009; Johnson et al., 2018; Matsuzaki et al., 2010; Saleh et al., 2019; Woo et al., 2012; Zuazo et al., 2019). This is evidenced by the recent clinical implementation of anti-PD-1 and anti-LAG-3 combinations (Burova et al., 2019; Chocarro et al., 2022b; Ghosh et al., 2019; Jiang et al., 2021; Sordo-Bahamonde et al., 2021; Tawbi et al., 2022; Zahm et al., 2021). Here, we carried out the most extensive and detailed study up-to-date on PD-1/LAG-3 co-expression as a signature of potent immune dysfunctionality in human cancers and T-cells. Overall, it included more than 12000 TCGA cancers and established the relationships between *PDCD1/LAG3* gene expression with immune cell infiltration and many biomarkers both in T-cells and within the tumour. The selection method led to a positive correlation between the *PDCD1/LAG3* signature with strong T-cell infiltration and specific pro-inflammatory immune cell types. This evidenced the association of the *PDCD1/LAG3* signature with potentially immunogenic “hot” cancers. This study encompassed almost 400 genes of T-cell function and activity, and correlations with more than 1000 regulators of cell cycle, cell-to-cell communication, chromatin organization, DNA repair, DNA replication, programmed cell death and autophagy. Our data also evidenced that T-cell infiltration could be characterised by an omics profile represented by PD-1/LAG-3/CTLA-4 immune checkpoints, or another one with TIM-3/TIGIT. Interestingly, GITR and VISTA showed independent patterns. Overall, these changes reflected pathways associated to high immune dysfunctionality and impaired T-cell function at all levels, including all functions related to the TCR signalosome, cytokine expression, inflammation, cell cycle, immunoproteasome, and signal transduction pathways among many others. Thus, the *PDCD1/LAG3* signature in tumours represented gene expression profiles characteristic of strong immune dysfunctionality, which included pathways regulating the activities of tumour-infiltrating T-cells.

Our data uncovered an exception with the *PDCD1/LAG3* profile in thymoma, which deviated noticeably. Thymoma cancers are rare and possess particular characteristics, such as inconsistent histological and biologic behaviour, without biomarkers that predict clinical behaviour (Kondo, 2008; Scorsetti et al., 2016). Currently, immune checkpoint blockade therapies are not standard of care for thymoma patients which seems to have increased rates of adverse events (Hao et al., 2022; Jakopovic et al., 2020), and our data suggest that PD-L1/PD-1 blockade or its combination with LAG-3 blockade may not work for thymoma.

Many of the dysfunctional features uncovered by multiomic studies were validated in T-cell lines with active PD-1, LAG-3 and PD-1+LAG-3 signalling pathways. The extensive T-cell phenotyping was consistent with PD-1 and LAG-3 signalling and was accentuated in T-cells with PD-1+LAG-3 co-signalling. Interestingly, we found only 35 proteins commonly regulated by PD-1, LAG-3 and PD- 1+LAG-3 co-signalling, but these were specially represented in all TCGA cancer types.

The engineering of PD-1+LAG-3 T-cell lines uncovered the full extent of T-cell dysfunctionality associated to this signature. For example, mTOR-AKT pathways were specifically up-regulated but several signalling pathways were also down-modulated by PD-1/LAG-3. Remarkably, we identified EIF2 signaling as a novel canonical pathway commonly suppressed by PD-1, LAG-3 and PD-1+LAG- 3 in T-cells. This is a relevant finding, because EIF2 pathway inhibition strongly impacts on translation, transcription, cellular stress responses and intracellular calcium signalling. These features are in agreement with the establishment of a strong T-cell dysfunctionality programme associated to PD- 1/LAG-3. To the best of our knowledge, no reports on the association between PD-1 or LAG-3 with the EIF2 pathway have been found up-to-date.

The E3 ubiquitin pathway of the CBL family was found to regulate EIF2 and TCR signalling by PD- 1/LAG-3. CBL proteins are known negative regulators of T-cell functions (Bachmaier et al., 2000; Chiang et al., 2000; Clemens, 2001; Dikic et al., 2003; Karwacz et al., 2011; Shamim et al., 2007). Our omics analyses have confirmed previous known features of E3 CBL proteins but in the context of PD- 1/LAG-3 co-signalling. For example, downmodulation of the MAPK and EIF2 signalling pathways, supported by previous studies that proposed CBL-B and C-CBL as negative regulators of MAPKs (Kales et al., 2014; Qu et al., 2011; Zhang et al., 2004; Zhao et al., 2013). Negative regulation of the MAPK pathway would disrupt phosphorylation of eukaryotic translation initiation factor, resulting in inhibition of EIF2 signalling (Siraj et al., 2022; Tian et al., 2011; Wu et al., 2022). Notably, although PD-1 has been broadly linked to CBL expression in T-cells (Babu et al., 2006; Karwacz et al., 2011; Kumar et al., 2021; Nguyen et al., 2021; Peer et al., 2017), LAG-3 expression has never been associated to CBL ubiquitin ligases before. Thus, we found that CBL-B and C-CBL were differentially regulated by PD-1, LAG-3 and PD-1+LAG-3 by co-blockade studies in primary T-cells from NSCLC patients.

Only co-blockade with a bispecific molecule simultaneously blocked both CBL-B and C-CBL expression. Hence, PD-1/LAG-3 co-blockade with CB213 showed a differential mechanism compared to just the combination of saturating concentrations of mono-specific anti-PD-1 and anti-LAG-3 antibodies. These results strongly indicate that PD-1/LAG-3 molecular complexes do have a biological meaning distinct than just the expression of PD-1 and LAG-3 separately. Therefore, a selective co-blockade strategy within LAG-3/PD-1 double-positive T-cells could benefit patients with intrinsic resistance to conventional immunotherapies. Further exploration of the molecular pathways uncovered in this study will probably lead to novel therapeutic targets, or help identifying a signature of response that could be used clinically.

## Data Availability

Mass-spectrometry data and search results files were deposited in the Proteome Xchange Consortium via the JPOST partner repository (https://repository.jpostdb.org; Reference: https://www.ncbi.nlm.nih.gov/pmc/articles/PMC5210561/) with the identifier PXD040408 for ProteomeXchange and JPST002062 for jPOST (for reviewers: https://repository.jpostdb.org/preview/118930299263fc6310a6249; Access key: 7060).

## Acknowledgments

We sincerely acknowledge all the patients and families that generously agreed to participate in this study. We are also thankful to the nursing staff of the Medical Oncology Day Care at University Hospital of Navarre who kindly provided the clinical samples. This research was supported by: The Spanish Association against Cancer (AECC), PROYE16001ESCO; Instituto de Salud Carlos III (ISCIII)-FEDER Project grants FIS PI20/00010, COV20/00000, and TRANSPOCART ICI19/00069; Biomedicine Project Grant from the Department of Health of the Government of Navarre-FEDER funds (BMED 050-2019, 51-2021); Strategic projects from the Department of Industry, Government of Navarre (AGATA, Ref. 0011-1411-2020-000013; LINTERNA, Ref. 0011-1411-2020-000033; DESCARTHES, 0011-1411-2019-000058); European Union Horizon 2020 ISOLDA project, under grant agreement ID: 848166. L.C. is financed by Instituto de Salud Carlos III (ISCIII), co-financed by FEDER funds, "Contratos PFIS: contratos predoctorales de formación en investigación en salud" (FI21/00080); M.E. is financed by the Navarrabiomed- Fundación Miguel Servet predoctoral contract.

## Author contributions

L.C., G.K. and D.E., conceptualised and designed the study; L.C., L.F-R., M.J.G-G. and D.E. performed cell lines engineering; L.C., L.F-R. and D.E. performed flow cytometric data acquisition and analysis; H.A. and R.V., recruited patients and supplied clinical samples; M.Z., A.B., H.A. and D.E. performed clinical samples processing and analysis; C.J., C.J.E., J.L., A.J.P., D.E. developed the CB213 Humabody; K.A., E.S. and J.F-R. generated the MS/MS library and performed the SWATH-MS-based experiments; L.C. and D.E. performed the bioinformatical and computational analysis of public databases; L.C., E.S., J.F-R, G.K. and D.E. performed the statistical analysis and interpretation of proteomic data; L.C., L.F-R., M.J.G-G., E.B., A.B., M.E., M.G., M.Z., H.A., K.A., E.S., J.F-I., G.K. and D.E. provided research support for performing experiments and data collection; L.C. and D.E. wrote the manuscript; G.K. and D.E. supervised the study. All authors critically reviewed the manuscript and approved its final version.

## Data availability

Mass-spectrometry data and search results files were deposited in the Proteome Xchange Consortium via the JPOST partner repository (https://repository.jpostdb.org; Reference: https://www.ncbi.nlm.nih.gov/pmc/articles/PMC5210561/) with the identifier PXD040408 for ProteomeXchange and JPST002062 for jPOST (for reviewers: https://repository.jpostdb.org/preview/118930299263fc6310a6249 ; Access key: 7060).

## MATERIALS AND METHODS

### Patients and clinical samples

Blood samples were prospectively collected from a cohort of patients with locally advanced or metastatic NSCLC treated with IC inhibitors (ICIs, nivolumab, pembrolizumab, atezolizumab) following current guidelines (Herbst et al., 2016; Horn et al., 2017; Rittmeyer et al., 2017) at University Hospital of Navarre between September 2017 and May 2019. The prospective observational study was approved by the Ethics Committee of Clinical Investigations at the University Hospital of Navarre (reference number: PI_2020/115). Informed consent was obtained from all subjects, and all experiments conformed to the principles in the WMA Declaration of Helsinki and the Department of Health and Human Services Belmont Report. Clinical details and composition of the cohort under study are described elsewhere (Arasanz et al., 2020; Bocanegra et al., 2022; Zuazo et al., 2019). Briefly, eligible patients were 18 years of age or older who had progressed to first-line platinum-based chemotherapy or concurrent chemoradiotherapy. A CT scan before the beginning of immunotherapy and another one after the first cycle of immunotherapy were performed. Four ml of peripheral blood samples were obtained immediately prior to the infusion of the first cycle of immunotherapy.

Responses were assessed following standard protocols according to current clinical practice based on RECIST 1.1 (Eisenhauer et al., 2009) and Immune-Related Response Criteria (Wolchok et al., 2009).

### Cells and lentivectors

Jurkat T cells were purchased from the ATCC, and cultured in standard conditions in RPMI medium supplemented with 10% FCS, penicillin/streptomycin and glutamine. Jurkat T cell lines were obtained by lentivector transduction and selection with puromycin, blasticidin or both in complete RPMI medium. Jurkat T cell lines were tested regularly (at least once a month) for mycoplasma by PCR. Human embryonic kidney (HEK) 293T cells were purchased from the ATCC and grown in DMEM supplemented with 10% FCS, penicillin/streptomycin and glutamine. 293T cells were routinely PCR- tested for mycoplasma.

Patient or healthy donor peripheral blood mononuclear cells (PBMCs) were isolated by FICOL gradients immediately after the blood extraction. PBMCs were washed and T cells were isolated as described (Zuazo et al., 2019). T cells were maintained in TexMACs medium (Miltenyi) until use.

The engineering and culture of SC3-A549 cells is described elsewhere (Zuazo et al., 2019). Briefly, these cells are human lung adenocarcinoma cells modified to express an anti-CD3 single chain antibody to stimulate T cells. Co-cultures of SC3-A549 cells with primary T cells were carried out as described at a 2:1 ratio (Zuazo et al., 2019). These cell cultures were performed in the presence of immune checkpoint inhibitors. PD-1, LAG-3, control isotype antibodies and their working concentrations used for the assays are described in (Zuazo et al., 2019). The bispecific Humabody CB213 that blocks simultaneously PD-1 and LAG-3 together with its control and their working concentrations are described in (Edwards et al., 2022) Co-cultures were carried out for two days before analyses.

### Plasmids and lentivector production

The coding sequences for LAG-3 and PD-1 genes were retrieved from https://www.ncbi.nlm.nih.gov/gene and https://www.uniprot.org/. Their cDNA sequences were synthetized (Geneart, Thermofisher) flanked by BamHI and NotI sites. Their extracellular stem, transmembrane and intracellular domains were amplified by PCR and fused to the extracellular domain of the SC3 construct in lentivectors as described before (Zuazo et al., 2019). Briefly, this molecule is a single-chain antibody derived from the anti-CD3 antibody OKT3 which is fused in frame to a human IgG transmembrane domain cloned under the transcriptional control of the SFFV promoter in a pDUAL-lentivector. For the constructs containing GFP or mCherry, their coding sequences were fused in frame to the intracellular signalling domains of PD-1 or LAG-3. All constructs were sequenced All the fusion genes were cloned into pDUAL-PuroR or pDUAL-BlastR lentivectors under the transcriptional control of the SFFV promoter. pDUAL-PuroR or pDUAL-BlastR lentivectors express puromycin or blasticidin resistance under the transcriptional control of the human ubiquitin promoter, and these are described in (Gato-Canas et al., 2017). To engineer lentivectors containing T-cell specific promoters, the promoter sequences for the following genes were retrieved from EPD Eukaryiotic Promoter Database (http://epd.vital-it.ch/human/human_database.php): *EF-1A*, *SNW1*, *HMGB1*, *WIBF*, *RPL13*, *RBM28*, *PDCD11*, *MICAL1*, *DDX21*, *SNRP70*, *ACAT2*, *HMGCS1*. Two kilobases upstream of the transcription start site were synthetized for each promoter (Geneart, Thermofisher), flanked by EcoRI and AscI restriction sites. Then, the SFFV promoter from the pSIN-GFP lentivector (Escors et al., 2008) was replaced by each of the T-cell specific promoters.

Lentivector production and titration were carried out as described elsewhere (Karwacz et al., 2011; Liechtenstein et al., 2014; Selden et al., 2007). Jurkat T-cells were transduced with a multiplicity of 10 and selected with puromycin (Gibco), blasticidin (Gibco) or both as described (Gato-Canas et al., 2017). Transduced cells were analysed for the expression of the target of interest by flow cytometry or by fluorescence microscopy. BioTeck Citation™ 5 cell imaging multi-mode reader-based fluorescence detection was used.

### Flow cytometry

Surface and intracellular stainings were carried out as described (Zuazo et al., 2019). The antibodies used were: CD4-APC-Vio770 (clone M-T466, Miltenyi Biotec), CD3-APC (clone REA613, Milenyi Biotec), CD28-PECy7 (clone CD28.2, Biolegend), PD-1-PE (clone EH12.2H7, Biolegend), CD8- FITC (clone SDK1, Biolegend), LAG3-PE (clone 11C3C65, Biolegend), LAG3-PerCP-Cy5.5 (clone 11C3C65, Biolegend), CD4-FITC (clone REA623, Milteny), CD62L-APC (clone 145/15, Milteny), CD3-PerCP-Cy5.5 (clone T100, TONBO), CD57-vioblue (clone TB03, Milteny), C3AR-PE (clone hC3aRZ8, Biolegend), CD25-PE (clone BC96, TONBO), CD39-APC-Cy7 (clone A1, Biolegend), CD158-PerCP-Cy5.5 (clone HP-MA4, Biolegend), CD16-PE-Vio770 (clone REA423, Milteny), CD137-PE (clone 4B4-1, Biolegend), CD69-PE (clone FN50, Biolegend), CD73-PE-Cy7 (clone AD2, Biolegend), HLA-DR-APC (clone REA332, Milteny), KLRG1-APC-Vio770 (clone REA261, Milteny), CD4-PE-Vio770 (clone REA261, Milteny), CD154-PerCP-Cy5.5 (clone 24-3I, Biolegend), CD223-PerCP-Cy5 (clone 11C3C65, Biolegend), TIM3-APC (clone F38-2E2, Biolegend), PD-1- Pacific Blue (clone EH12.2H7, Biolegend). When required, surface staining was followed by intracellular staining using the BD Transcription Factor buffer set or the BD Fixation/permeabilization Kit. The following antibodies were used for intracellular staining: anti-Ki67-APC or pacific blue-conjugated antibodies (Clone ki67, Biolegend), AF648-CBL-B (G1 clone, Santa Cruz Biotech), H2AX-FITC (1:100 dilution, clone 2F3, reference 613403, BioLegend),AF648-C-CBL (A-9 clone, Santa Cruz Biotech), IFN-γ-FITC (clone REA600, Milteny), IFN-α/β (clone FAB245P, RD Systems), IL-12/13 (clone C11.5, Biolegend), IL4 (clone REA895, Milteny), IL-2-APC (clone 17H12, Biolegend), IL17A-BV421 (clone BL168, Biolegend), TNF-α (clone Mab11, Biolegend) and IL-10 (clone REA842, Milteny) antibodies. Flow cytometry was carried out with a BD FACS CANTO flow cytometer. Data was analysed by Flowjo.

### Proteomics and data analysis

Cell proteomes were analysed by SWATH-MS (Collins et al., 2017). Cell pellets were homogenized in a lysis buffer containing 7 M urea, 2 M thiourea, and 50 mM DTT. The homogenates were spun down at 100,000 g for 1 h at 15°C. Protein quantitation was performed by Bradford (Bio-Rad). Protein in-solution digestion, peptide purification, and reconstitution prior to mass spectrometric analysis and library generation were performed as previously reported (Ferrer et al., 2021).

Peptides recovered from in-gel digestion processing were reconstituted into a final concentration of 0.5 μg/μL of 2% ACN, 0.5% FA, 97.5% Milli-Q-water prior to mass spectrometric analysis. MS/MS data sets for spectral library generation were acquired on a Triple TOF 5600+ mass spectrometer (Sciex, Canada) interfaced to an Eksigent nanoLC ultra 2D pump system (SCIEX, Canada) fitted with a 75 μm ID column (Thermo Scientific 0.075 × 250 mm, particle size 3 μm and pore size 100 Å). Prior to separation, the peptides were concentrated on a C18 precolumn (Thermo Scientific 0.1 × 50 mm, particle size 5 μm and pore size 100 Å). Mobile phases were 100% water 0.1% formic acid (FA) (buffer A) and 100% Acetonitrile 0.1% FA (buffer B). Column gradient was developed in a gradient from 2% B to 40% B in 120 min. Column was equilibrated in 95% B for 10 min and 2% B for 10 min. During all processes, the precolumn was in line with column and flow was maintained all along the gradient at 300 nL/min. Output of the separation column was directly coupled to nanoelectrospray source. MS1 spectra was collected in the range of 350−1250 m/z for 250 ms. The 35 most intense precursors with charge states of 2 to 5 that exceeded 150 counts per second were selected for fragmentation, rolling collision energy was used for fragmentation, and MS2 spectra were collected in the range of 230−1500 m/z for 100 ms. The precursor ions were dynamically excluded from reselection for 15 s. MS/MS data acquisition was performed using AnalystTF 1.7 (Sciex) and spectra files were processed through ProteinPilot v5.0 search engine (Sciex) using Paragon Algorithm (v.4.0.0.0) (Shilov et al., 2007) for database search. To avoid using the same spectral evidence in more than one protein, the identified proteins were grouped based on MS/MS spectra by the Progroup algorithm, regardless of the peptide sequence assigned. The protein within each group that could explain more spectral data with confidence was depicted as the primary protein of the group. False discovery rate was performed using a nonlinear fitting method (Tang et al., 2008)and displayed results were those reporting a 1% Global false discovery rate or better.

For SWATH-MS-based experiments, the instrument (Sciex Triple- TOF 5600+) was configured as described elsewhere (Gillet et al., 2012). Briefly, the mass spectrometer was operated using an isolation width of 16 Da (15 Da of optimal ion transmission efficiency and 1 Da for the window overlap), a set of 37 overlapping windows were constructed covering the mass range 450−1000 Da. In this way, 1 μL of ach sample was loaded onto a trap column (Thermo Scientific 0.1 × 50 mm, particle size 5 μm and pore size 100 Å) and desalted with 0.1% TFA at 3 μL/ min during 10 min. The peptides were loaded onto an analytical column (Thermo Scientific 0.075 × 250 mm, particle size 3 μm and pore size 100 Å) equilibrated in 2% acetonitrile 0.1% FA. Peptide elution was carried out with a linear gradient of 2 to 40% B in 120 min (mobile phases A:100% water 0.1% formic acid (FA) and B: 100% Acetonitrile 0.1% FA) at a flow rate of 300 nL/min. Eluted peptides were infused in the mass spectrometer. The Triple-TOF was operated in swath mode, in which a 0.050 s TOF MS scan from 350 to 1250 m/z was performed, followed by 0.080 s product ion scans from 230 to 1800 m/z on the 37 defined windows (3.05 s/cycle). Collision energy was set to optimum energy for a 2 + ion at the center of each SWATH block with a 15 eV collision energy spread. The mass spectrometer was always operated in high sensitivity mode. The resulting ProteinPilot group file from library generation was loaded into PeakView (v2.1, Sciex) and peaks from SWATH runs were extracted with a peptide confidence threshold of 99% confidence (Unused Score ≥1.3) and a false discovery rate (FDR) lower than 1%. For this, the MS/MS spectra of the assigned peptides was extracted by ProteinPilot, and only the proteins that fulfilled the following criteria were validated: (1) peptide mass tolerance lower than 10 ppm, (2) 99% of confidence level in peptide identification, and (3) complete b/ y ions series found in the MS/MS spectrum. Only proteins quantified with at least two unique peptides were considered. The quantitative data obtained by PeakView were analysed using Perseus software (Tyanova et al., 2016) for statistical analysis and data visualization. Output files with the identified proteins were then managed with Perseus for subsequent statistical analysis. An unpaired Student t-test was used for direct comparisons between two groups of samples. Differential PD-1/LAG-3 proteins versus the SC3 control condition comparisons were identified, following p-value ≤ 0.05, Log2 (Fold Change) ≥ 0.38 and Log2 (Fold Change) ≤ -0.38 criteria.

### Bioinformatics, computational and statistical analyses

Public transcriptomic and genomic databases for TCGA human cancers were analysed with TIMER 2.0 (Li et al., 2016; Li et al., 2017; Li et al., 2020), GEPIA (Gene Expression Profiling Interactive Analysis) (Tang et al., 2019; Tang et al., 2017), cBioportal (Cerami et al., 2012; Gao et al., 2013), TNM Plot (Bartha and Gyorffy, 2021), and Ingenuity Pathway Analysis (IPA) (Kramer et al., 2014).

TIMER 2.0 uses six algorithms grouped into the R package IMMUNEDECONV for transcriptome analyses in immunooncology (Li et al., 2020; Sturm et al., 2019). Briefly, IMMUNEDECONV integrates TIMER, xCell, MCP-counter, CIBERSORT, EPIC and quanTIseq to provide a highly accurate estimation of immune cell infiltration. TIMER was previously developed (Li et al., 2017) to take into account tissue specificity and distinct immune contexts to estimate immune infiltration for 6 cell types (Pao et al., 2018). XCell produces estimations on a higher number of immune cell types but without considering tumour purity as a confounding factor (Aran et al., 2017). MCP-counter produces estimations of immune cell and stromal cell infiltration (Becht et al., 2016). CIBERSORT deconvolves from transcriptomic data more detailed T-cell signatures to provide infiltration estimates of many T-cell subsets (Newman et al., 2015). EPIC (Racle et al., 2017) and quanTIseq (Finotello et al., 2019) generate scores for cell fractions. TIMER 2.0 generates a table of Spearmańs correlations between input genes and infiltration/abundance of 59 immune cell types and subtypes (Li et al., 2020). The correlation with tumour purity can also be performed as well as the association of gene expression with cell type.

GEPIA performs differential transcriptional gene expression quantification, calculates survival and prognostic maps and similar-gene analyses. GEPIA utilizes the UCSC Xena data of TCGA and GTEx (Vivian et al., 2017) which contains 198619 coding transcripts which include isoforms grouped with non-coding transcripts. This allows the focused analyses on 84 cancer subtypes to provide a curated list of signatures such as tumour microenvironment and infiltrating immune content in the form of a signature score.

Multidimensional cancer genomics data was additionally analysed using cBIOPORTAL. These analyses include alterations such as gene frequencies, translocations and duplications within a single study or across multiple studies. The portal also supports biological pathway exploration, and calculation of survival plots. Genomic data includes somatic mutations, DNA copy number alternations, RNA expression patterns (including mRNA and microRNA), DNA methylation, protein and phosphoprotein abundance. Ten cancer studies and 20 TCGA studies are used to extract the dataset in cBIOPORTAL (Barretina et al., 2012; Barretina et al., 2010; Cancer Genome Atlas, 2012a, b; Cancer Genome Atlas Research, 2008, 2011, 2012; Taylor et al., 2010) Comparisons of gene expression between healthy tissue, tumour and metastatic tumour were carried out with TNMplot from NCBIGEO, TCGA, TARGET and GTEx repositories, which encompass a total dataset of 56938 samples. Gene expression comparisons were performed by non-parametric tests such as Mann-Whitney or Kruskal-Wallis.

Differential protein expression data from proteomic analyses by PERSEUS were analysed with Ingenuity Pathway Analysis (IPA) (Kramer et al., 2014), Metascape (Zhou et al., 2019), Reactome (Croft et al.), STRING (Szklarczyk et al., 2019) and TFactS (Essaghir and Demoulin, 2012; Essaghir et al., 2010). IPA compares a dataset of genes to the Ingenuity Knowledge Base and applies four causal analytic algorithms to identify upstream regulators, mechanistic networks, causal network analysis and downstream effects analysis. IPA utilizes two scores for inference; P-values from a Fisheŕs exact test to obtain an enrichment score, and a Z-score to assess the match of observed and predicted regulation patterns. The specific IPA settings were used for the analyses:

Dataset upload, ID/Observation name: UniProt/Swiss-Prot Accession; Measurement/Annotation: Expr Log Ratio. Analysis, Analyse/filter dataset: Core analysis; Based on this dataset, core analysis type “Expression analysis”; Measurement type, Expr Long Ratio, pre-analysis filtering.

### General Settings

Ingenuity Knowledge base reference set considered for p-value calculations. Direct and indirect relationships were considered for networks and upstream regulator analysis.

### Networks

Maximum of 25 networks, where each network can contain up to 35 molecules. Causal networks were generated, where score was calculated using causal paths only.

### Node types

Biological drugs, canonical pathways, chemical (- endogenous mammalian, endogenous non- mammalian, -kinase inhibitor, -other, -protease inhibitor), chemical drugs, chemical reagent, chemical toxicant, complex, cytokine, disease, enzyme, function, fusion gene/ product, G-protein coupled receptor, group, growth factor, ion channel, kinase, ligand-dependent nuclear receptor, mature microRNA, microRNA, peptidase, phosphatase, transcription regulator, translation regulator, transmembrane receptor, transporter and others were included.

### Data sources

Ingenuity expert information: Ingenuity expert findings and ingenuity expert assist findings; Ingenuity supported third party information: microRNA-mRNA interactions (miRecords, TarBase, TargetScan Human, TargetScan Mouse); Protein-protein interactions: BIND, Cognia, DIP, Interactome studies, MIPS; Additional Sources: An Open Access Database of Genome-wide Association Results, BioGRID, Catalogue of Somatic Mutations in Cancer (COSMIC), Chemical Carcinogenesis Research Information System (CCRIS), Clinical Genome Resource (ClinGen), ClinicalTrials.gov, GVK Biosciences, Hazardous Substances Data Bank (HSDB), HumanCyc, IntAct, miRbase, Mouse Genome Database (MGD), Obesity Gene Map Database, Online Mendelian Inheritance in Man (OMIM).

### miRNA confidence

Experimentally observed miRNA confidence was considered:

### Species

Mammal (human, mouse, rat) and uncategorized species with an stringent filter (filter molecules and relationships) to include only molecules and relationships from the Ingenuity Knowledge Base that are pertinent to the selected species

### Tissues and cell lines

Tissues and Primary Cells; Tissues and Primary Cells not otherwise specified; Cells: cells not otherwise specified, adypocytes, astrocytes, beta islet cells, blood patelets, bone marrow cells (bone marrow cells not othersie specified, megakaryocytes, other bone marrow cells), cardiomyocites, chrondrocytes, enthotelial cells (endothelial cells not otherwise specified, HUVEC cells, microvascular endothelial cells, other endothelial cells), epithelial cells (epithelial cells not otherwise specified, hepatocytes, keratynocytes, melanocytes, Sertoli cells, other epithelial cells), fibroblasts, granulosa cells, hematopoietic progenitor cells, immune cells (immune cells not otherwise specified, dendritic cells (dendritic cells not otherwise specified, BDCA-1+ dendritic cells, BDCDA-3+ dendritic cells, bone marrow-derived dendritic cells, CD34+ cells, Langerhans cells, monocyte-derived dendritic cells (monocyte-derived dendritic cells not otherwise specified, immature monocyte-derived dendritic cells, mature monocyte-derived dendritic cells, other monocyte-derived dendritic cells), myeloid dendritic cells, plasmacytoid dendritic cells, other dendritic cells), granulocytes (granulocytes not otherwise specified, eosinophils, neutrophils, other granulocytes)), neurons (neurons not otherwise specified, cortical neurons, granule cells, purkinje cells, pyramidal neurons, other neurons), oocytes, osteoblasts, smooth muscle cells (smooth muscle cells not otherwise specified, vascular smooth muscle cells, other smooth muscle cells), splenocytes, stem cells (stem cells not otherwise specified, embryonic stem cells, mesenchymal stem cells, other stem cells), stromal cells, other cells); Nervous system: nervous system not otherwise specified, amygdala, brain, brainstem, caudate nucleus, cerecellum, cerebral cortex, cerebral ventricles, chroroid plexus, corpus callosum, dorsal room ganglion, granule cell layer, gray matter, hippocampus, hypothalamus, medulla oblongata, nucleus accumbens, olfactory bulb, parietal lobe, pituitary gland, putamen, sciatic nerve, spinal cord, striatum, substantia nigra, subventricular zone, thalamus, trigeminal ganglion, ventricular zone, white matter, other nervous system; Organ system: organ system not otherwise specified, adipose, adrenal gland, bladder, calvaria, cartilage tissue, cornea, crypt, dermis, epidermis, esophagus, forestomach, heart, kidney, large intestine, lens, liver, lung, lymph node, mammary gland, ovary, pancreas, placenta, prostate gland, retina, salivary gland, skeletal muscle, spleen, stomach, tests, thymus, thyroid gland, trachea, uterus, other organ systems,; Other tissues and primary cells; Cell line; Cell line not otherwise specified:

Breast cancer cell lines: breast cancer cell lines not otherwise specified, BT-474, BT549, HS578T, MCF7, MDA-MB-231, MDA-MB-361, MDA-MB-435, MDA-MB-468, MDA-N, NCI-ADR-RES, T47-D, other breast cancer cell lines. Cervical cancer cell line: cervical cancer cell line not otherwise specified, HeLa, other cervical cancer cell line. CNS Cell lines: CNS cell lines not otherwise specifid, SF-2687, SF-295, SF-539, SNB-75, U251, U87MG, other CNS cell lines. Colon cancer cell lines: colon cancer cell lines not otherwise specified, Caco2 cells, COLO205, HCC-2998, HCT-116, HCT- 15, HT29, KM-12, RKO, SW-E480, SW-620, other colon cancer cell lines. Fibroblast cell lines: fibroblast cell lines not otherwise specified, 3T3-L1 cells, Cos-7 cells, MEF cells, NIH/3T3 cells, swiss 3T3 cells, other fibroblast cell lines. Hepatoma cell lines: hepatoma cell lines not otherwise specified, Hep3B, HepG2, HyH7, other hepatoma cell lines. Immune cell lines: immune cell lines not otherwise specified, BA/F3, HMC-1, J774, RBL-2H3, other immune cell lines. Kidney cancer cell lines: kidney cancer cell lines not otherwise specified, 786-0, A498, ACHN, CAKI-1, RXF-393, SN12C, TK-10, UO-31. Kidney cell lines: kidney cell lines not otherwise specified, 293 cells, other kidney cell lines.

Leukemia cell lines: leukemia cell lines not otherwise specified, CCRF-CEM, HEL, HL-60, Jurkat, K- 562, MOLT-4, NB4, THP-1, other leukemia cell lines. Lung Cancer cell lines: lung cancer cell lines not otherswise specified, A549-ATCC, EKVX, H460, HOP-62, HOP-92, NCI-H226, NCI-H23, NCI-H332M, NCI-H522, other lung cancer cell lines. Lymphoma cell lines: lymphoma cell lines not otherwise specified, SR, U937, WEHI-231, other lymphoma cell lines. Macrophage cancer cell lines: macrophage cancer cell lines not otherwise specified, J-774A.1, RAW 264-7, other macrophage cancer cell lines. Melanoma cell lines: melanoma cell lines not otherwise specified, A375, LOX IMVI, M14, MALME-3M, SK-MEL-2, SK-MEL-28, UACC-257, UACC-62, other melanoma cell lines. Myeloma cell lines: myeloma cell lines not otherwise specified, RPMI-8266, U266, other myeloma cell lines. Neuroblastoma cell lines: neuroblastoma cell lines not otherwise specified, SK-N-SH, other neuroblastoma cell lines. Osteosarcoma cell lines: osteosarcoma cell lines not otherwise specified, MG- 63, U2OS, other osteosarcoma cell lines. Ovarian cancer cell lines: ovarian cancer cell lines not otherwise specified, A27780, IGROV1, OVCAR-3, OVCAR-4, OVCAR-5, OVCAR-8, SK-OV-3, other ovarian cancer cell lines. Pancreatic cancer cell lines: pancreatic cancer cell lines not otherwise specified, INS-1, Min6, PANC-1, other pancreatic cancer cell lines. Pheochromocytoma cell lines: pheochromocytoma cell lines not otherwise specified, PC-12 cells, other pheochromocytoma cell lines. Prostate cancer cell lines: prostate cancer cell lines not otherwise specified, DU-145, LNCaP cells, PC- 3, other prostate cancer cell lines. Teratocarcinoma cell lines. Other cell lines: The filter category "Other" contains findings for specific tissues or cell lines that do not have their own specific category below the parent cell line or tissue type grouping. The filter categories "Tissue and Primary Cells not otherwise specified" and "Cell Line not otherwise specified" contain findings that do not have any additional detailed categorization beyond the parent cell line or tissue type grouping name.

### Mutations

Functional effect: gain-of-function, knockout, loss of function, null mutation; Inheritance mode: dominant, recessive, X-linked, Y-linked; Translation impact: frameshift, in-frame, missense, nonsense, silent; Unclassified mutation; Zygosity: hemizygous, heterozygous, homozygous; Wild type Mutation-based relationships were included in the analysis. Mutation findings from the QIAGEN Knowledge Base are available for inclusion in the networks. Such findings involve a mutant form of at least one of the genes.

Set cutoffs; Dataset column: Expr Log Ratio; Measurement Value type: Expr Long Ratio; Range: -1.0 to 1.0

Metascape combines OMICs studies to analyse and interpret the data by searching in 40 independent databases which allows comparative analyses across multiple experiments. Metascape utilizes hypergeometric tests and Benjamini-Hochberg p-value correction to identify ontology terms within an input list. Metascape uses Gene Ontology (Ashburner et al., 2000), KEGG (Kanehisa and Goto, 2000), Reactome and MSigDB (Subramanian et al., 2005) for enrichment analyses, pairwise-tested with a Kappa test. STRING was used to complement IPA and Metascape to infer functional interactomes using the proteomics dataset, which uses available public sources of protein-protein interactions.

Additional statistical analyses of data from flow cytometry studies were performed with GraphPad 8 as previously described (Gato-Canas et al., 2017; Karwacz et al., 2011; Zuazo et al., 2019). Variables under study were subjected to Kolmogorov-Smirnov normality tests to ensure the use of parametric tests. Percentages of cell marker expression by flow cytometry were normally distributed. In these cases, parametric tests were used using two-way ANOVA tests followed by Tukey’s pair-wise comparisons for multi-comparison studies. In these ANOVA tests, the independent experiments were considered a random factor to remove inter-experimental variation. Progression-free and overall survivals were represented by Kaplan-Meier plots, and statistical significance tested with Log-Rank tests. Experiments on T-cell phenotyping were independently repeated a minimum of three times, each repetition corresponding to biological replicates and not technical replicates. No data was considered an outlier.

## KEY ABRREVIATIONS

• PD-1: “Programmed cell death protein 1” protein.
• LAG-3: “Lymphocyte-activation gene 3” protein.
• *PDCD1* or *PD1*: “Programmed cell death protein 1” gene.
• *LAG3*: “Lymphocyte-activation gene 3” gene.
• *CTLA4*: “Cytotoxic T-Lymphocyte Antigen 4” gene.
• *TIM3* or *HAVCR2*: “Hepatitis A Virus Cellular Receptor 2” gene, also known as “T-cell immunoglobulin and mucin domain-containing protein 3 (TIM3)” gene.
• *TIGIT*: “T-Cell Immunoreceptor With Ig And ITIM Domains” gene.
• *GITR* or *TNFRSF18*: “Tumour necrosis factor receptor superfamily member 18” gene, also known as “glucocorticoid-induced TNFR-related protein (GITR)” gene.
• *VISTA*, *C10ORF54* or *VSIR*: “V-Set Immunoregulatory Receptor”, also known as “V-Type Immunoglobulin Domain-Containing Suppressor Of T-Cell Activation” gene.
• *PDCD1/LAG3* profile or signature: we refer as *PDCD1/LAG3* in this manuscript as the gene expression profile commonly associated to *PDCD1* and *LAG3* expression profiles.
• PD-1/LAG-3 profile or signature: we refer as PD-1/LAG-3 in this manuscript as the protein expression profile commonly associated to PD-1 and LAG-3 expression profiles.
• *PDCD1* and *LAG3* profiles or signatures: we refer as *LAG3* and *PDCD1* in this manuscript as the sum of *LAG3* gene profile and *PDCD1* gene profile separately.
• PD-1 and LAG-3 profiles or signatures: we refer as PD-1 and LAG-3 in this manuscript as the sum of PD-1 protein profile and LAG-3 protein profile separately.
• PD-1+LAG-3: we refer as PD-1+LAG-3 in this manuscript as the modified T-cell lines co-expressing SC3-PD-1 and SC3-LAG-3 molecules.
• TCR: T-cell receptor.
• *TOX*: “Thymocyte selection-associated high mobility group box factor” gene.

