## Supplemental tables and figures for "PD-1/LAG-3 Dysfunctionality Signatures in Human Cancers"

| Condition | *Overall survival, median time (months) (95% CI)* |
|---:|---|
| *LAG3* | 50.50 (38.20 - 63.91) |
| *PDCD1* | 90.11 (66.74 - NA) |
| *CTLA4* | 71.01 (35.54 - NA) |
| *TIM3* | 90.87 (89.33 - NA) |
| *TIGIT* | 78.18 (45.83 - NA) |
| *GITR* | 106.95 (51.29 - NA) |
| *LAG3*, *PDCD1* | 49.02 (38.89 - NA) |
| *LAG3*, *CTLA4* | NA |
| *LAG3*, *TIM3* | 43.43 (13.84 - NA) |
| *PDCD1*, *CTLA4* | 35.18 (17.16 - NA) |
| *LAG3, TIGIT* | NA |
| *PDCD1, TIM3* | NA |
| *LAG3, GITR* | 67.36 (17.92 - NA) |
| *PDCD1, TIGIT* | 90.38 (47.80 - NA) |
| *CTLA4, TIM3* | NA |
| *PDCD1, GITR* | 49.05 (13.58 - NA) |
| *CTLA4, TIGIT* | 45.14 (34.75 - NA) |
| *CTLA4, GITR* | NA |
| *TIM3, TIGIT* | 55.43 (55.43 - NA) |
| *TIM3, GITR* | NA |
| *TIGIT, GITR* | 93.83 (44.84 - NA) |
| *LAG3, PDCD1, TIM3* | NA |
| *LAG3, CTLA4, TIM3* | NA |
| *LAG3, PDCD1, GITR* | 11.38 |
| *PDCD1, CTLA4, TIM3* | NA |
| *PDCD1, CTLA4, TIGIT* | NA |
| *PDCD1, CTLA4, GITR* | 6.18 |
| *PDCD1, TIM3, TIGIT* | NA |
| *PDCD1, TIGIT, GITR* | 15.78 |
| *TIM3, TIGIT, GITR* | 8.35 |
| *LAG3, PDCD1, TIM3, TIGIT* | NA |

**Supplementary Table 1.** Correlation between overall survivals with the expression of the indicated immune checkpoints combinations in tumor samples.

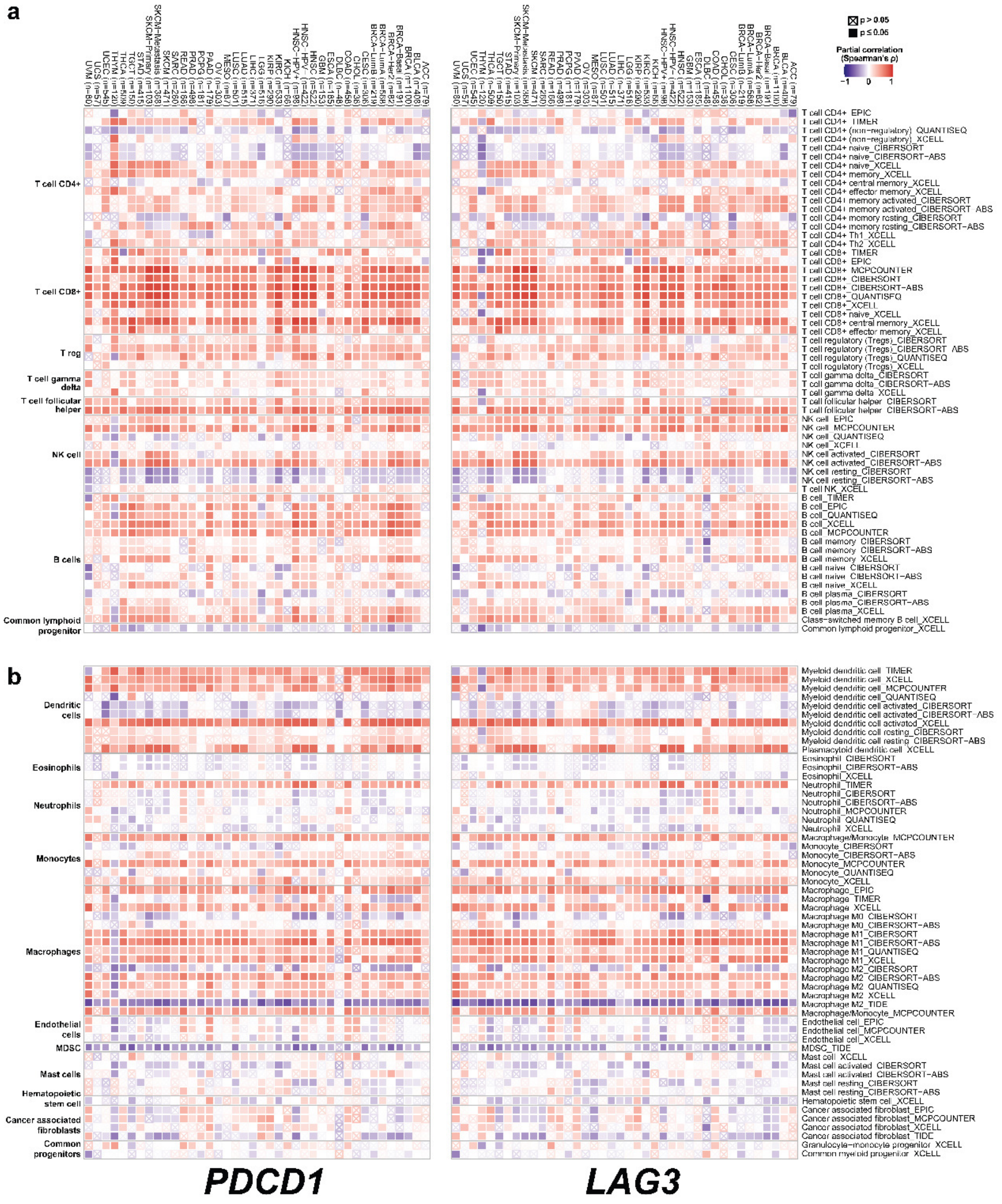

**Figure S1. Correlation of *PDCD1* and *LAG3* expression with immune cell infiltrates.** Heat maps represent correlation of immune cell infiltration data with *PDCD1* or *LAG3* expression in a total number of 12159 samples distributed on TCGA cancers. Statistical significance was tested with a partial purity-adjusted Spearman's correlation test. **(a)** Correlation with lymphoid infiltrates. **(b)** Correlation with non-lymphoid infiltrates. Specific immune cell types and their infiltration stage are indicated. Cancer types are specified on top. Red, positive correlation; blue, negative correlation. Crossed squares indicate no significant correlation (p>0.05).

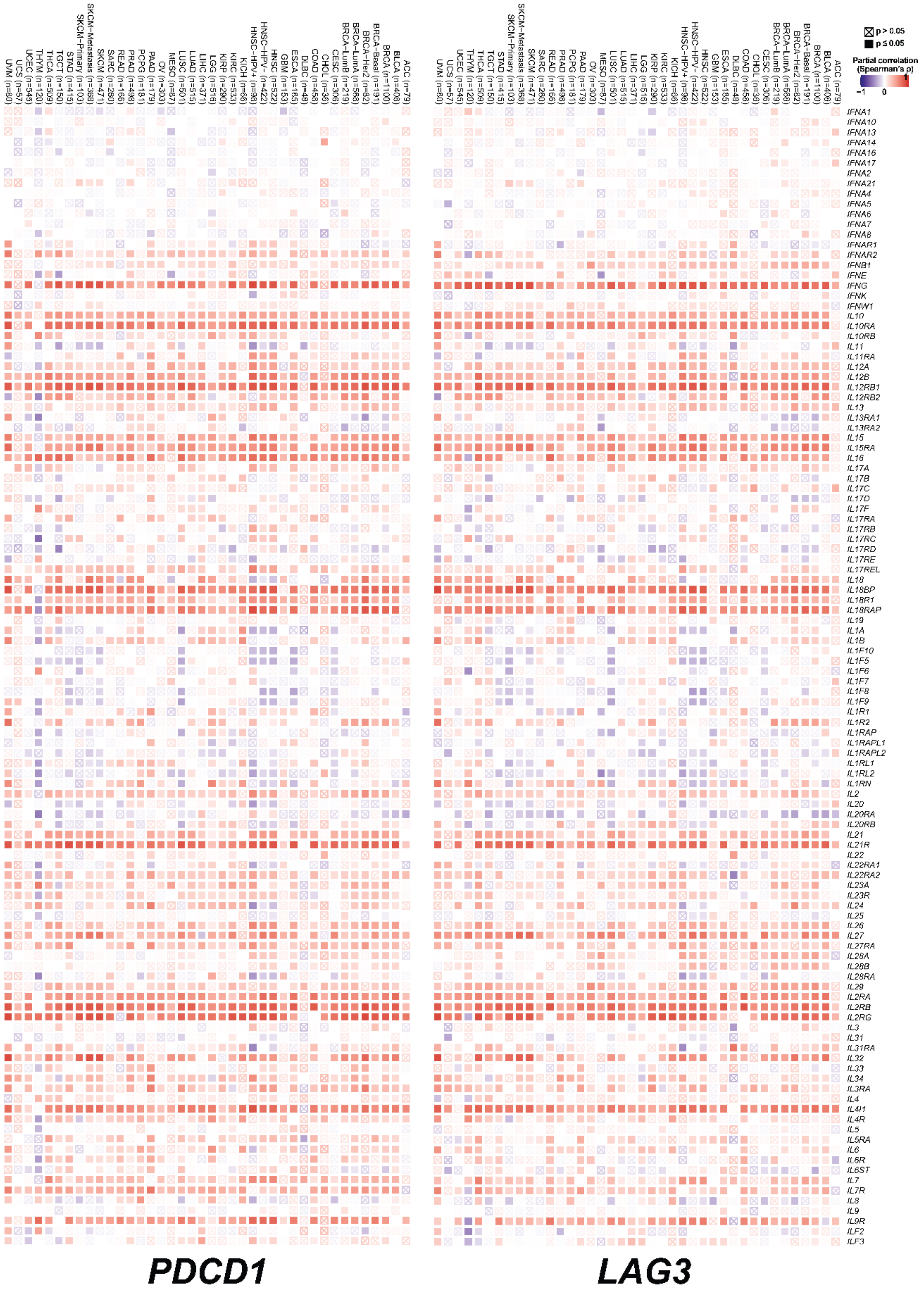

**Figure S2. Correlation of *PDCD1* and *LAG3* gene expression with IFN and IL gene expression in TCGA cancers (n=12159).** Heat maps show correlates of *PDCD1* and *LAG3* gene expression with the indicated genes (right), as shown for each cancer type (top). A partial purity-adjusted Spearman's correlation was used to test statistical significance. Red, positive correlation; blue, negative correlation. Crossed squares indicate no significant correlation (p>0.05).

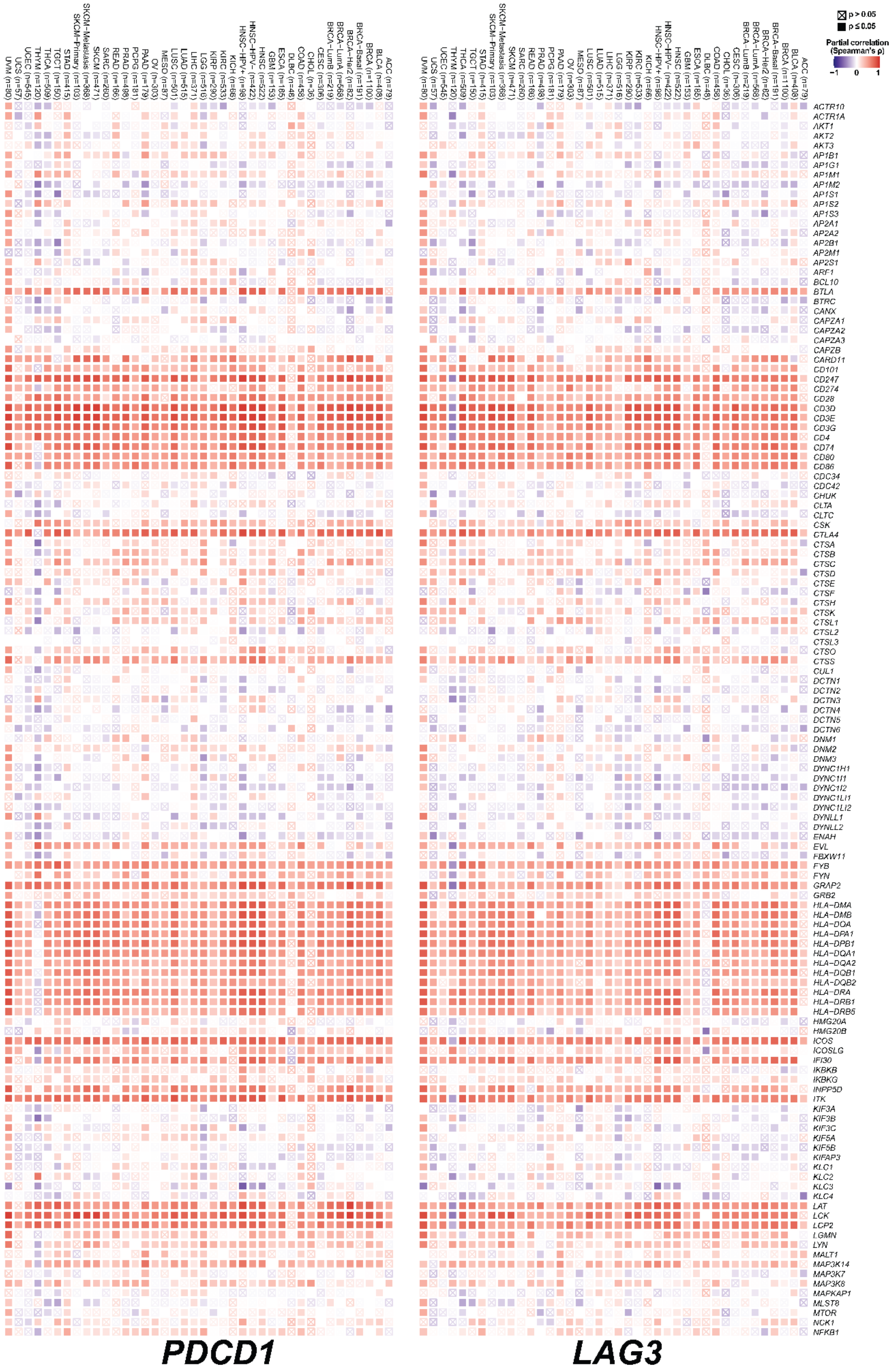
p > 0.05
p ≤ 0.05
Partial correlation (Spearman's ρ)
−1
0
1
ACTR10
ACTR1A
AKT1
AKT2
AKT3
AP1B1
AP1G1
AP1M1
AP1M2
AP1S1
AP1S2
AP1S3
AP2A1
AP2A2
AP2B1
AP2M1
AP2S1
ARF1
BCL10
BTLA
BTRC
CANX
CAPZA1
CAPZA2
CAPZA3
CAPZB
CARD11
CD101
CD247
CD274
CD28
CD3D
CD3E
CD3G
CD4
CD74
CD80
CD86
CDC34
CDC42
CHUK
CLTA
CLTC
CSK
CTLA4
CTSA
CTSB
CTSC
CTSD
CTSE
CTSF
CTSH
CTSK
CTSL1
CTSL2
CTSL3
CTSO
CTSS
CUL1
DCTN1
DCTN2
DCTN3
DCTN4
DCTN5
DCTN6
DNM1
DNM2
DNM3
DYNC1H1
DYNC1I1
DYNC1I2
DYNC1LI1
DYNC1LI2
DYNLL1
DYNLL2
ENAH
EVL
FBXW11
FYB
FYN
GRAP2
GRB2
HLA−DMA
HLA−DMB
HLA−DOA
HLA−DPA1
HLA−DPB1
HLA−DQA1
HLA−DQA2
HLA−DQB1
HLA−DQB2
HLA−DRA
HLA−DRB1
HLA−DRB5
HMG20A
HMG20B
ICOS
ICOSLG
IFI30
IKBKB
IKBKG
INPP5D
ITK
KIF3A
KIF3B
KIF3C
KIF5A
KIF5B
KIFAP3
KLC1
KLC2
KLC3
KLC4
LAT
LCK
LCP2
LGMN
LYN
MALT1
MAP3K14
MAP3K7
MAP3K8
MAPKAP1
MLST8
MTOR
NCK1
NFKB1
PDCD1
LAG3

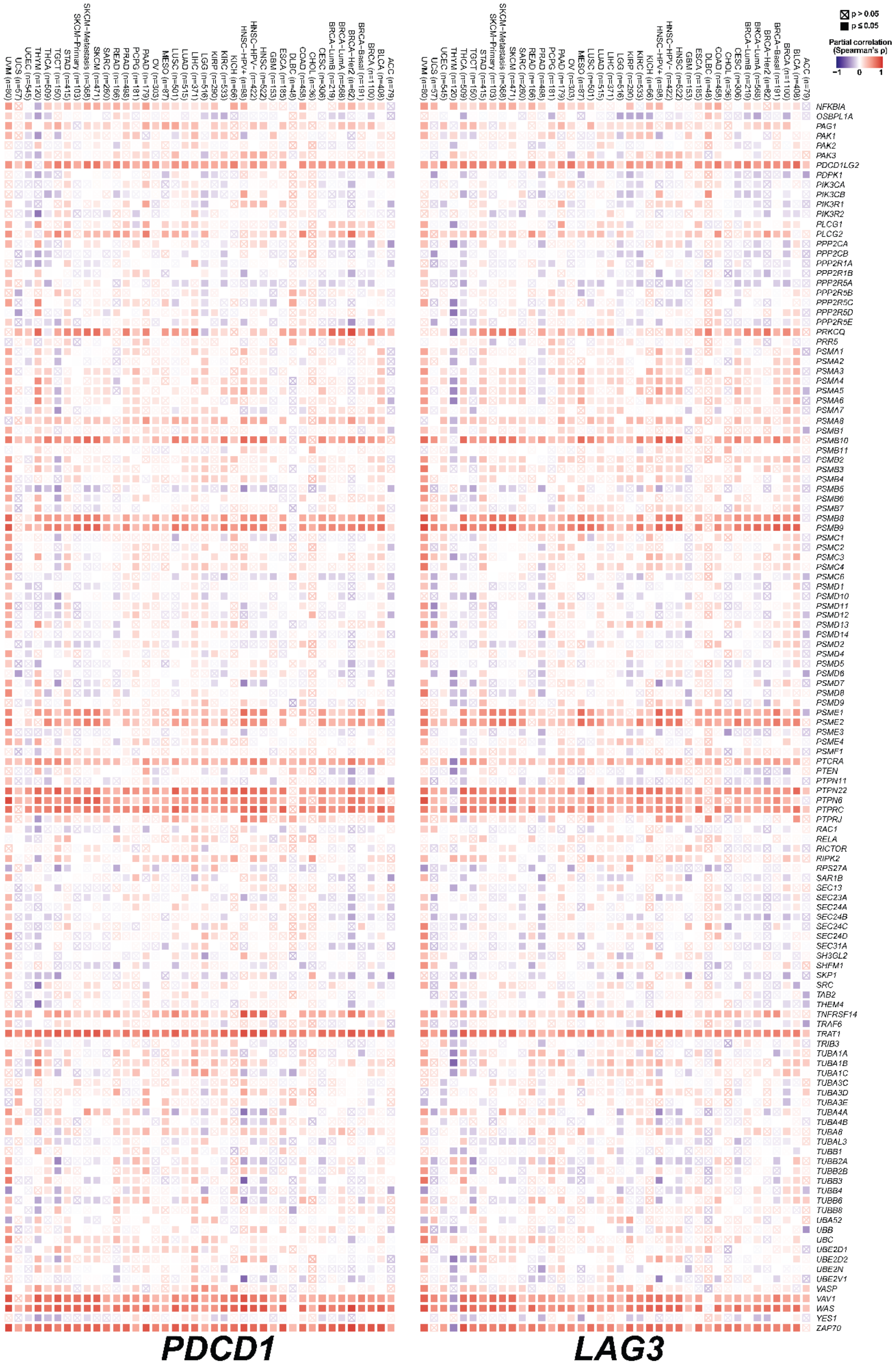

**Figure S3. Correlation of *PDCD1* and *LAG3* gene expression correlation with gene expression characteristic of TCR signalling, coestimulation by the CD28 family and**

**MHCII antigen presentation in TCGA cancers (n=12159).** Heat maps show correlates of *PDCD1* and *LAG3* gene expression with the indicated genes (right), as shown for each cancer type (top). A partial purity-adjusted Spearman's correlation was used to test statistical significance. Red, positive correlation; blue, negative correlation. Crossed squares indicate no significant correlation (p>0.05).

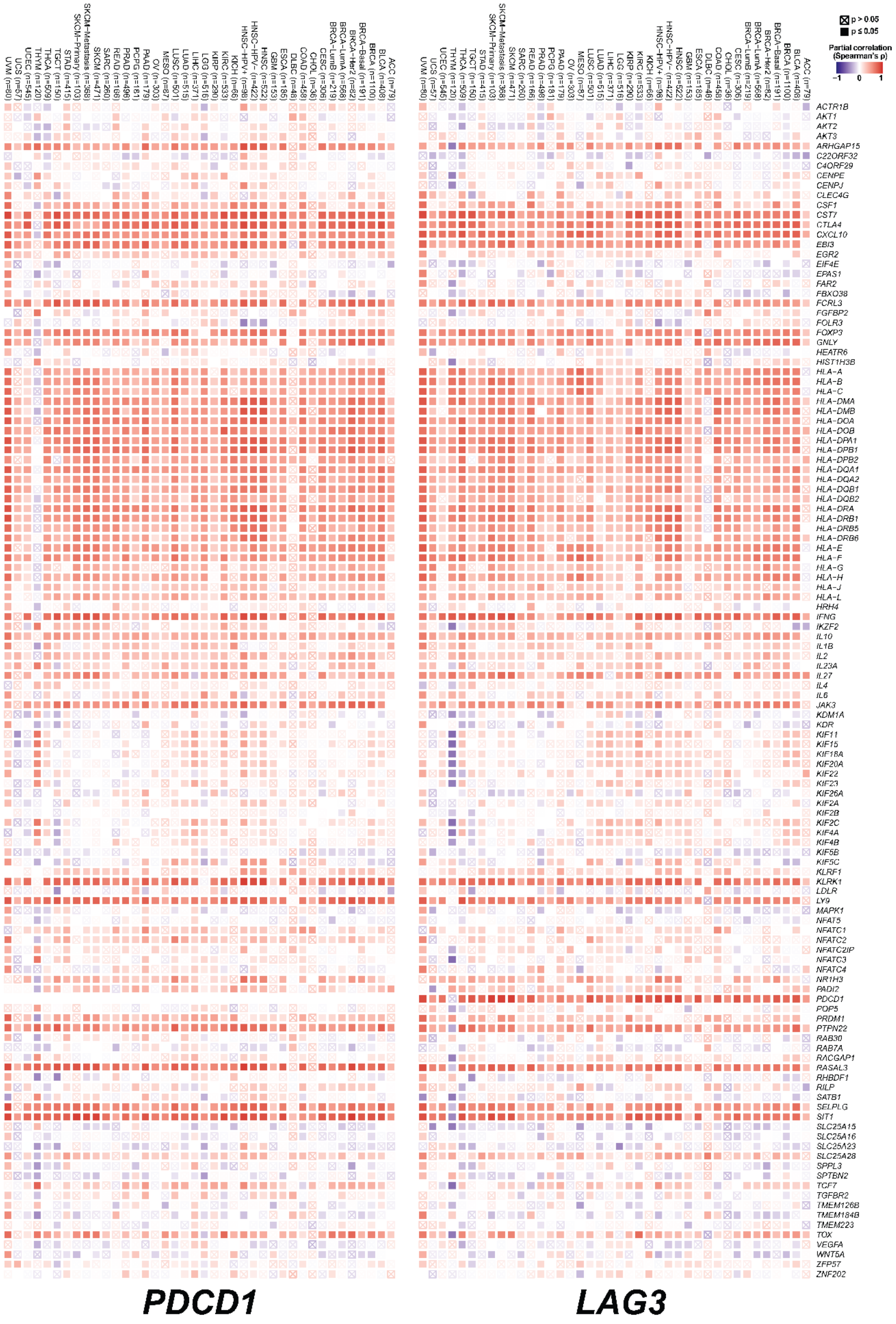


**Figure S4. Correlation of *PDCD1* and *LAG3* gene expression with IPA- predicted gene expression regulatory networks and causal relationships in TCGA cancers.** Heat maps show correlates of *PDCD1* and *LAG3* gene expression with the indicated genes, as shown for each cancer type (top). These molecules are associated with downstream targets which negatively regulated the TCR signalosome, including genes involved in immune synapse formation, TCR-associated kinases, phosphatases, cytokine signalling kinases and PI3K/AKT signalling pathway, among others, as shown in Figure 3. A partial purity-adjusted Spearman's correlation was used to test statistical significance. Red, positive correlation; blue, negative correlation. Crossed squares indicate no significant correlation ($p>0.05$). Cancer types are indicated on top, and specific targets are named on the right.

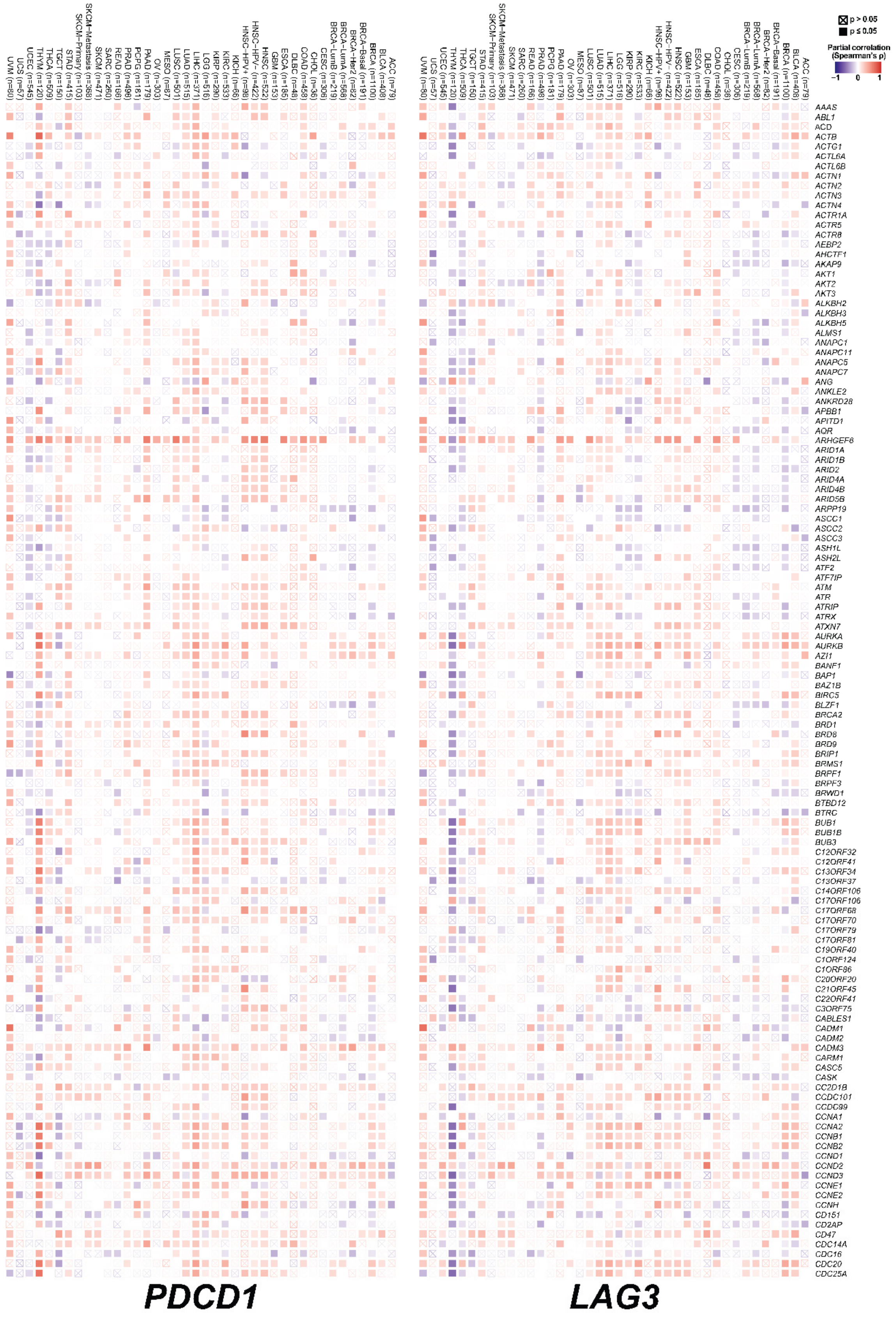
p > 0.05
p ≤ 0.05
Partial correlation (Spearman's ρ)
−1
0
1
UVM (n=80)
UCS (n=57)
UCEC (n=545)
THYM (n=120)
THCA (n=509)
TGCT (n=150)
STAD (n=415)
SKCM-Primary (n=103)
SKCM-Metastasis (n=368)
SKCM (n=471)
SARC (n=260)
READ (n=166)
PRAD (n=498)
PCPG (n=181)
PAAD (n=179)
OV (n=303)
MESO (n=87)
LUSC (n=501)
LUAD (n=515)
LIHC (n=371)
LGG (n=516)
KIRP (n=290)
KIRC (n=533)
KICH (n=66)
HNSC-HPV+ (n=98)
HNSC-HPV- (n=422)
HNSC (n=522)
GBM (n=153)
ESCA (n=185)
DLBC (n=48)
COAD (n=458)
CHOL (n=36)
CESC (n=306)
BRCA-LumB (n=219)
BRCA-LumA (n=568)
BRCA-Her2 (n=82)
BRCA-Basal (n=191)
BRCA (n=1100)
BLCA (n=408)
ACC (n=79)
AAAS
ABL1
ACD
ACTB
ACTG1
ACTL6A
ACTL6B
ACTN1
ACTN2
ACTN3
ACTN4
ACTR1A
ACTR5
ACTR8
AEBP2
AHCTF1
AKAP9
AKT1
AKT2
AKT3
ALKBH2
ALKBH3
ALKBH5
ALMS1
ANAPC1
ANAPC11
ANAPC5
ANAPC7
ANG
ANKLE2
ANKRD28
APBB1
APITD1
AQR
ARHGEF6
ARID1A
ARID1B
ARID2
ARID4A
ARID4B
ARID5B
ARPP19
ASCC1
ASCC2
ASCC3
ASH1L
ASH2L
ATF2
ATF7IP
ATM
ATR
ATRIP
ATRX
ATXN7
AURKA
AURKB
AZI1
BANF1
BAP1
BAZ1B
BIRC5
BLZF1
BRCA2
BRD1
BRD8
BRD9
BRIP1
BRMS1
BRPF1
BRPF3
BRWD1
BTBD12
BTRC
BUB1
BUB1B
BUB3
C12ORF32
C12ORF41
C13ORF34
C13ORF37
C14ORF106
C17ORF106
C17ORF68
C17ORF70
C17ORF79
C17ORF81
C19ORF40
C1ORF124
C1ORF86
C20ORF20
C21ORF45
C22ORF41
C3ORF75
CABLES1
CADM1
CADM2
CADM3
CARM1
CASC5
CASK
CC2D1B
CCDC101
CCDC99
CCNA1
CCNA2
CCNB1
CCNB2
CCND1
CCND2
CCND3
CCNE1
CCNE2
CCNH
CD151
CD2AP
CD47
CDC14A
CDC16
CDC20
CDC25A
PDCD1
LAG3

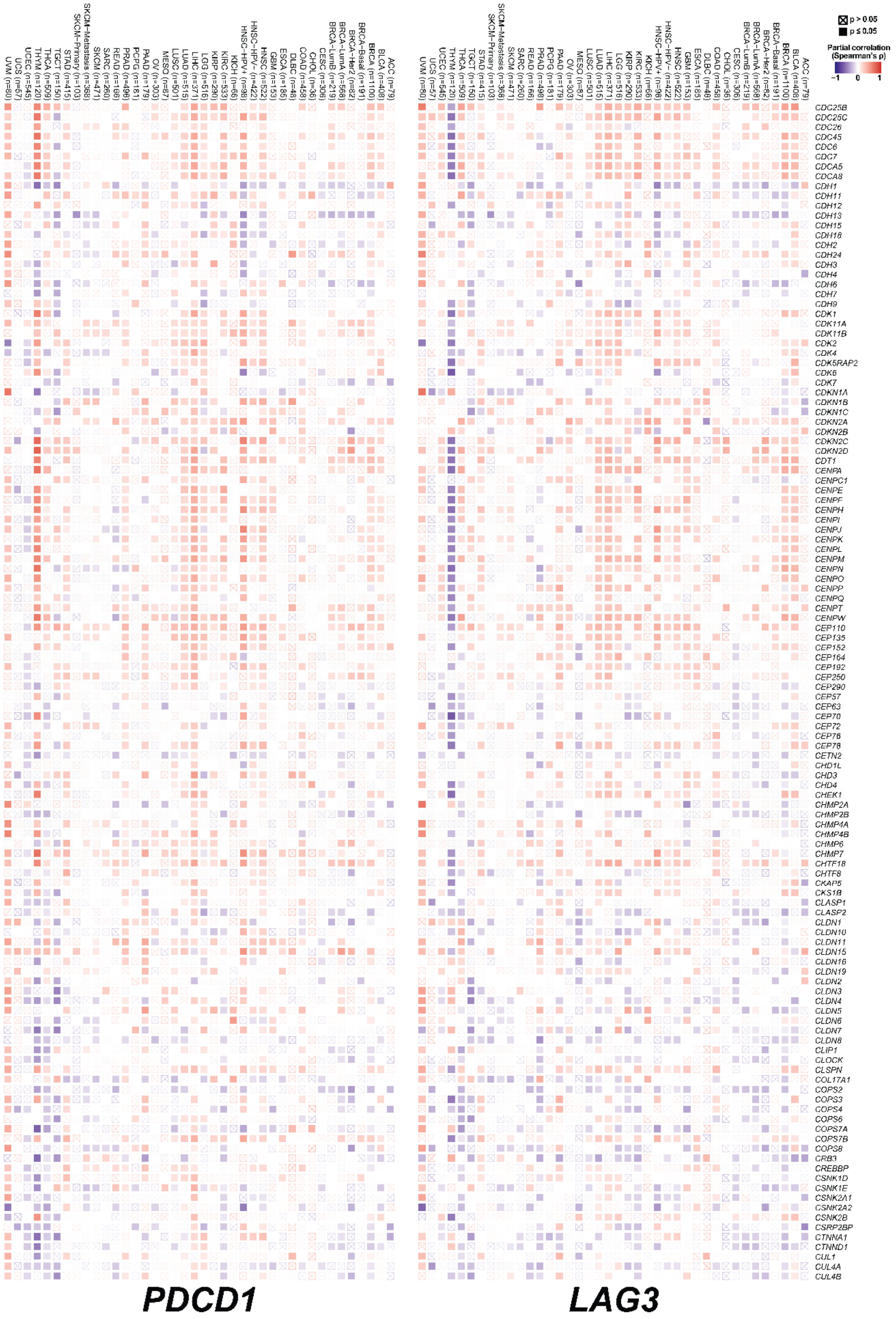
p > 0.05
p ≤ 0.05
Partial correlation (Spearman's ρ)
-1
0
1
PDCD1
LAG3

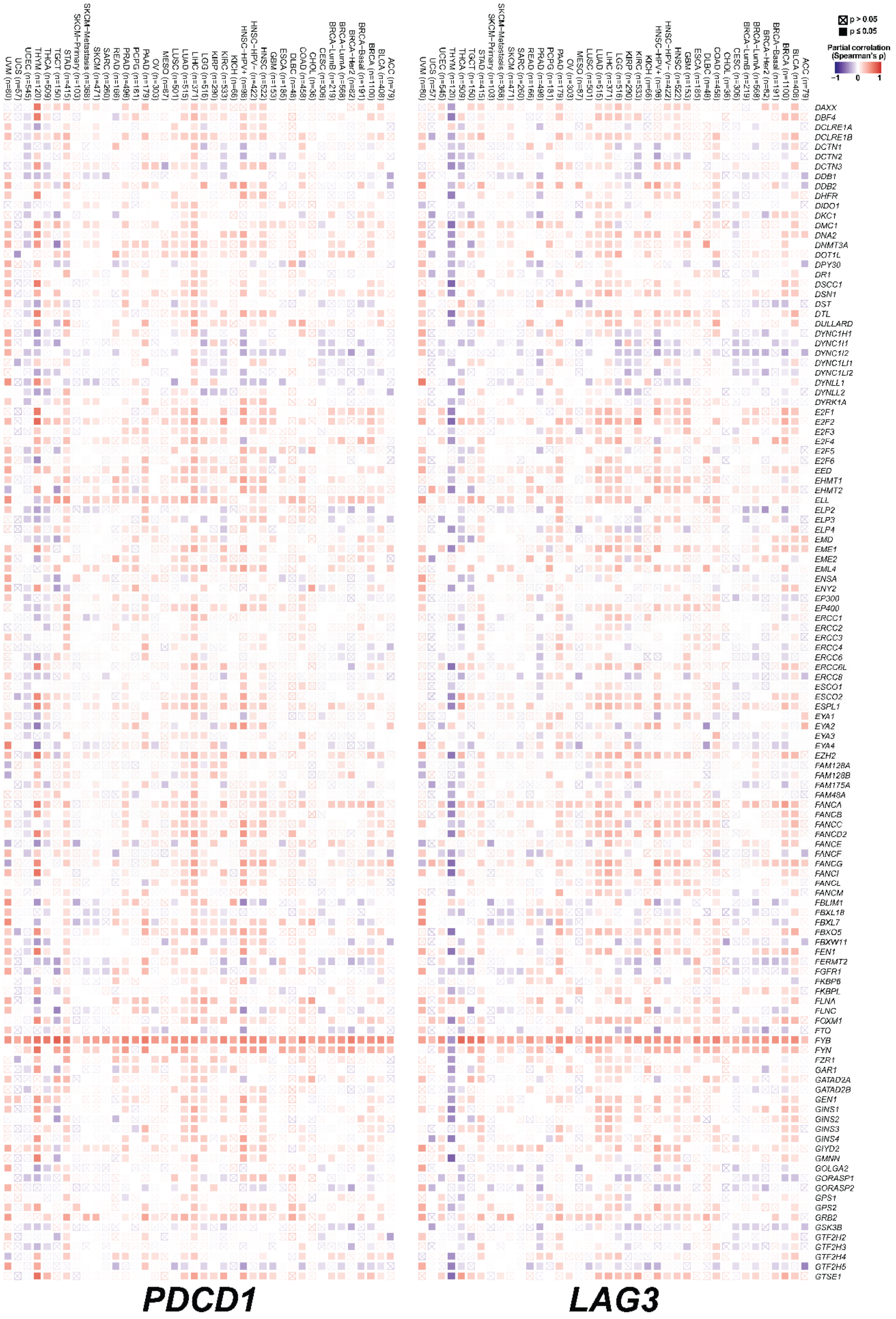
PDCD1
LAG3
p > 0.05
p ≤ 0.05
Partial correlation (Spearman's ρ)
−1
0
1

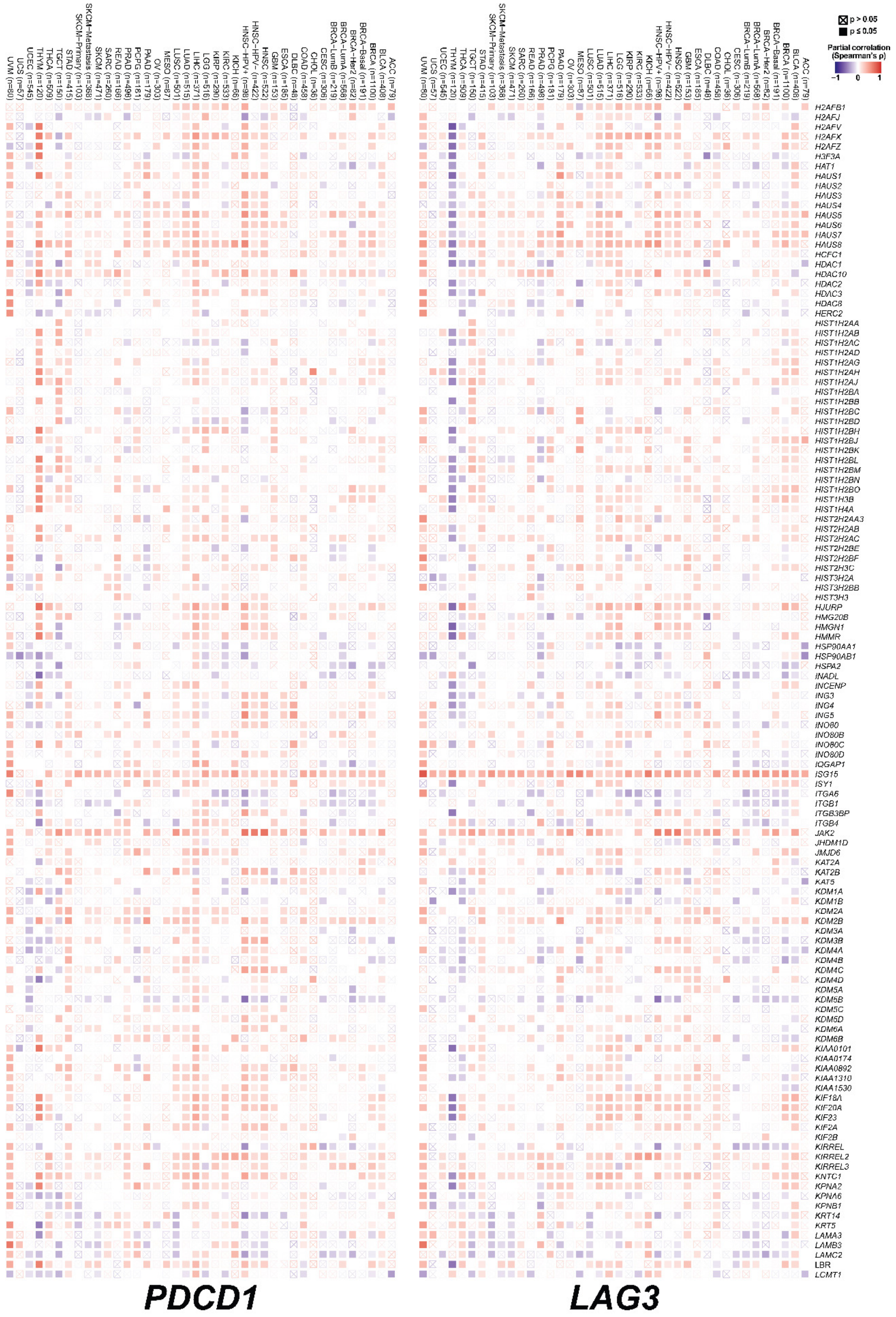
p > 0.05
p ≤ 0.05
Partial correlation (Spearman's ρ)
−1
0
1
UVM (n=80)
UCS (n=57)
UCEC (n=545)
THYM (n=120)
THCA (n=509)
TGCT (n=150)
STAD (n=415)
SKCM-Primary (n=103)
SKCM-Metastasis (n=368)
SKCM (n=471)
SARC (n=260)
READ (n=166)
PRAD (n=498)
PCPG (n=181)
PAAD (n=179)
OV (n=303)
MESO (n=87)
LUSC (n=501)
LUAD (n=515)
LIHC (n=371)
LGG (n=516)
KIRP (n=290)
KIRC (n=533)
KICH (n=66)
HNSC-HPV+ (n=98)
HNSC-HPV- (n=422)
HNSC (n=522)
GBM (n=153)
ESCA (n=185)
DLBC (n=48)
COAD (n=458)
CHOL (n=36)
CESC (n=306)
BRCA-LumB (n=219)
BRCA-LumA (n=568)
BRCA-Her2 (n=82)
BRCA-Basal (n=191)
BRCA (n=1100)
BLCA (n=408)
ACC (n=79)
H2AFB1
H2AFJ
H2AFV
H2AFX
H2AFZ
H3F3A
HAT1
HAUS1
HAUS2
HAUS3
HAUS4
HAUS5
HAUS6
HAUS7
HAUS8
HCFC1
HDAC1
HDAC10
HDAC2
HDAC3
HDAC8
HERC2
HIST1H2AA
HIST1H2AB
HIST1H2AC
HIST1H2AD
HIST1H2AG
HIST1H2AH
HIST1H2AJ
HIST1H2BA
HIST1H2BB
HIST1H2BC
HIST1H2BD
HIST1H2BH
HIST1H2BJ
HIST1H2BK
HIST1H2BL
HIST1H2BM
HIST1H2BN
HIST1H2BO
HIST1H3B
HIST1H4A
HIST2H2AA3
HIST2H2AB
HIST2H2AC
HIST2H2BE
HIST2H2BF
HIST2H3C
HIST3H2A
HIST3H2BB
HIST3H3
HJURP
HMG20B
HMGN1
HMMR
HSP90AA1
HSP90AB1
HSPA2
INADL
INCENP
ING3
ING4
ING5
INO80
INO80B
INO80C
INO80D
IQGAP1
ISG15
ISY1
ITGA6
ITGB1
ITGB3BP
ITGB4
JAK2
JHDM1D
JMJD6
KAT2A
KAT2B
KAT5
KDM1A
KDM1B
KDM2A
KDM2B
KDM3A
KDM3B
KDM4A
KDM4B
KDM4C
KDM4D
KDM5A
KDM5B
KDM5C
KDM5D
KDM6A
KDM6B
KIAA0101
KIAA0174
KIAA0892
KIAA1310
KIAA1530
KIF18A
KIF20A
KIF23
KIF2A
KIF2B
KIRREL
KIRREL2
KIRREL3
KNTC1
KPNA2
KPNA6
KPNB1
KRT14
KRT5
LAMA3
LAMB3
LAMC2
LBR
LCMT1
PDCD1
LAG3

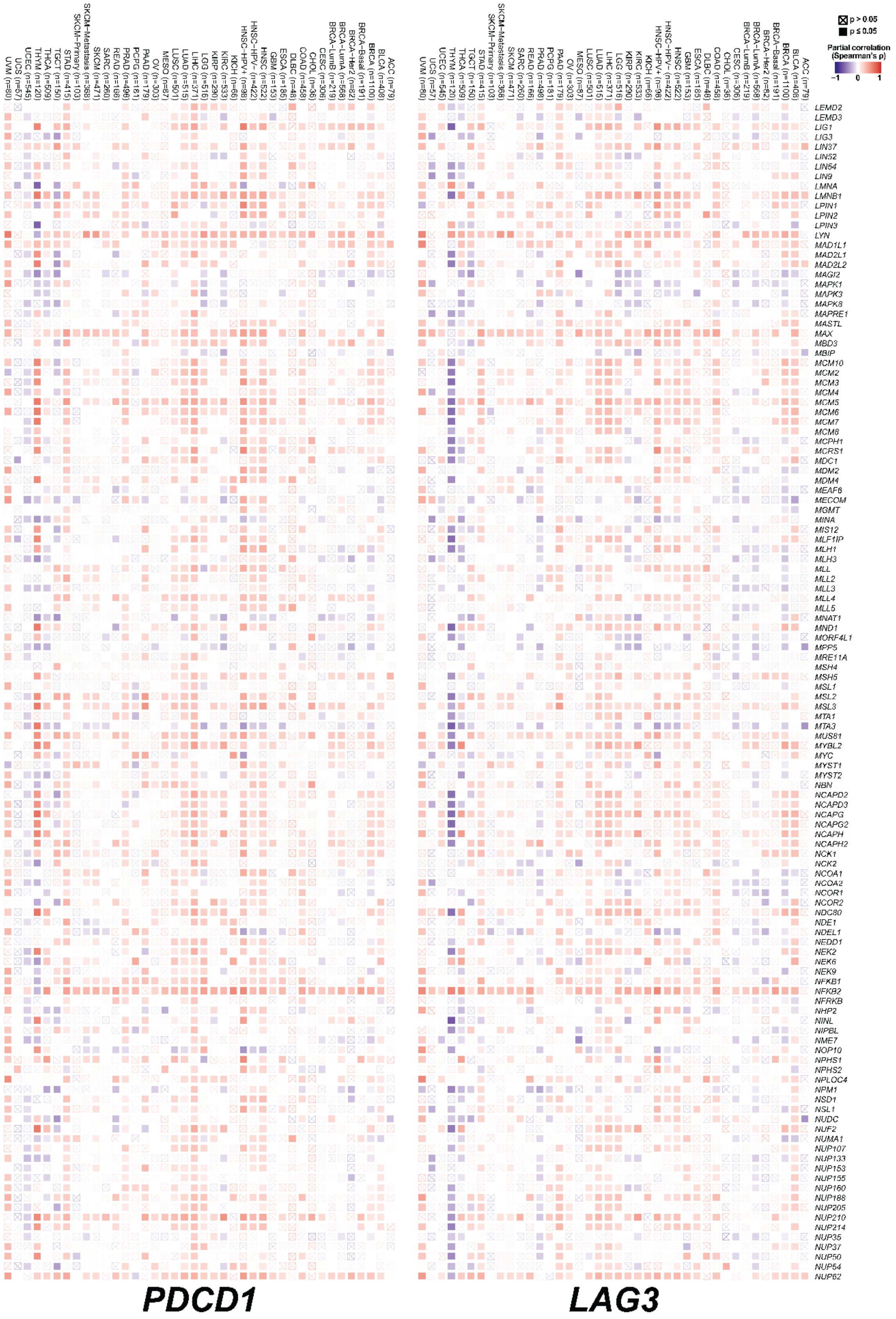

p > 0.05
p ≤ 0.05
Partial correlation (Spearman's ρ)
−1
0
1
PDCD1
LAG3

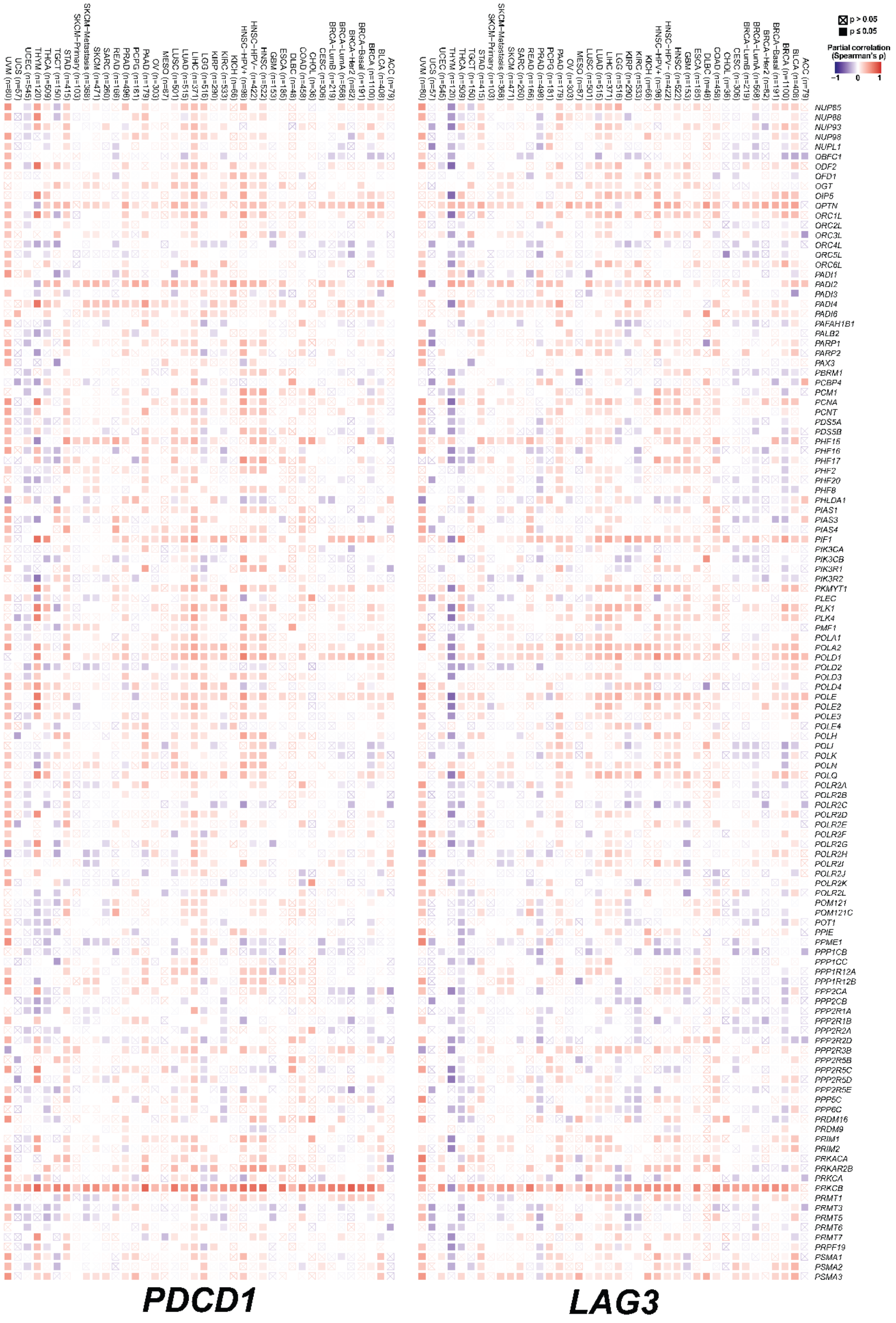
PDCD1
LAG3
p > 0.05
p ≤ 0.05
Partial correlation (Spearman's ρ)
-1
0
1

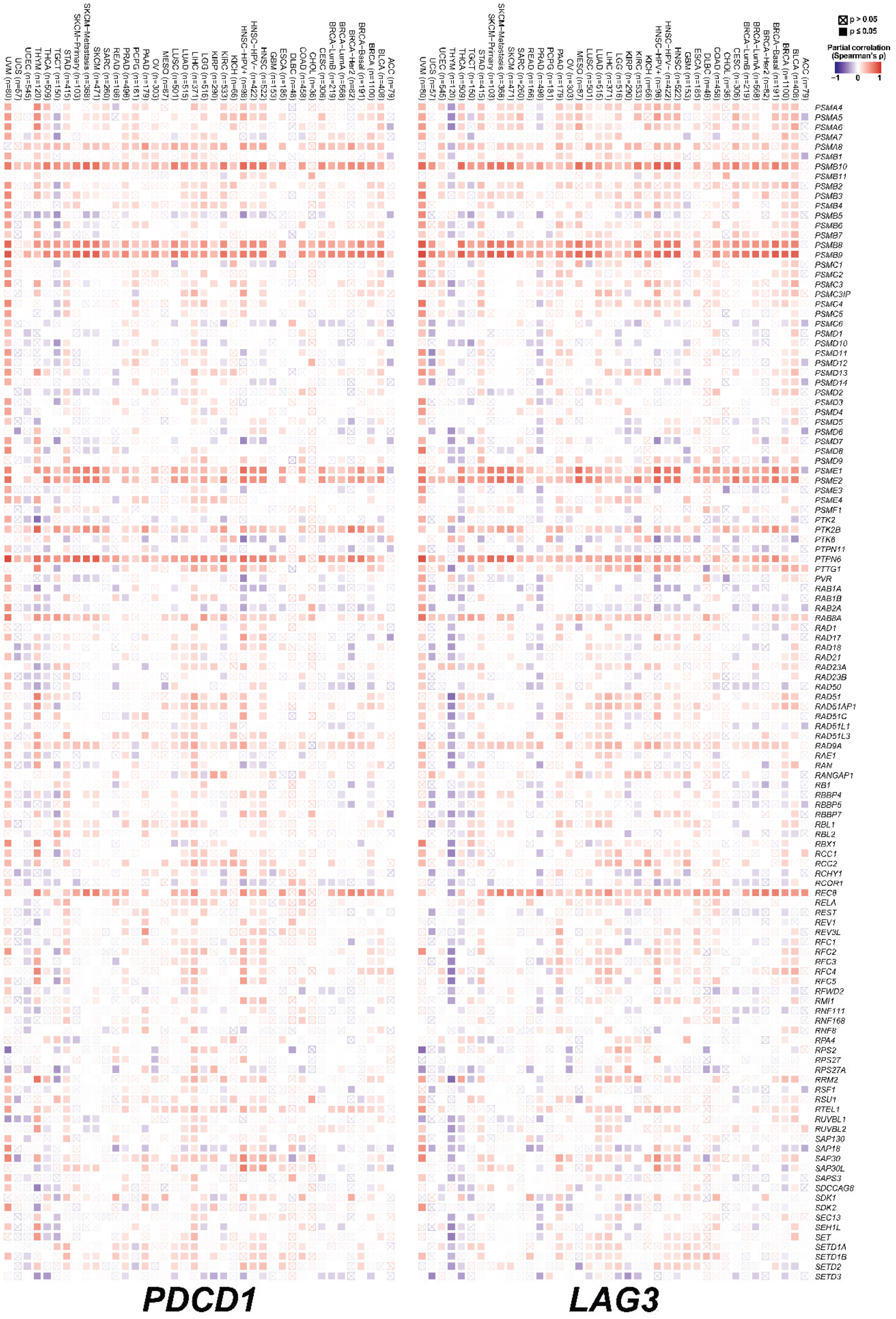
PDCD1
LAG3
p > 0.05
p ≤ 0.05
Partial correlation (Spearman's ρ)
−1
0
1

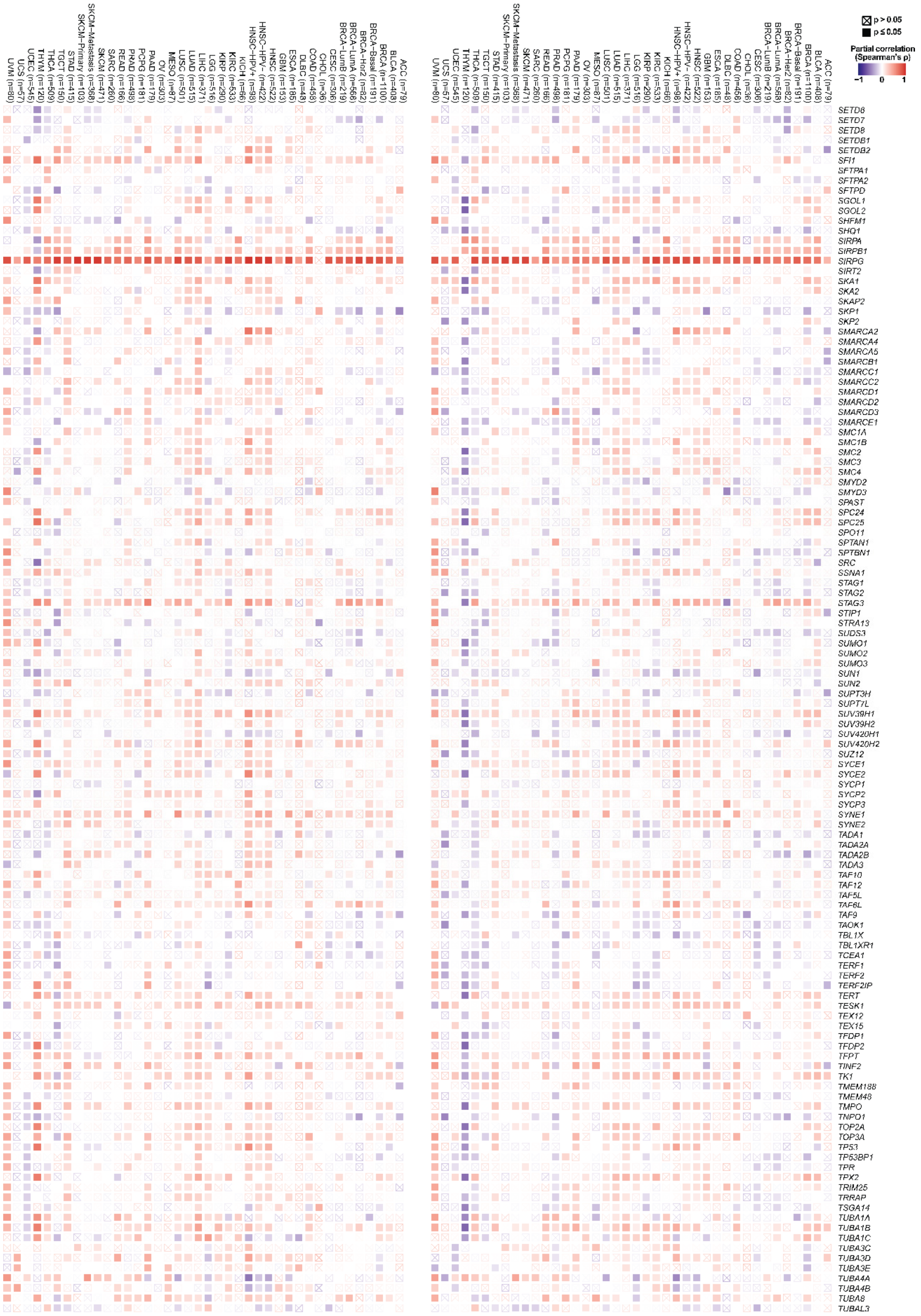

**PDCD1** **LAG3**

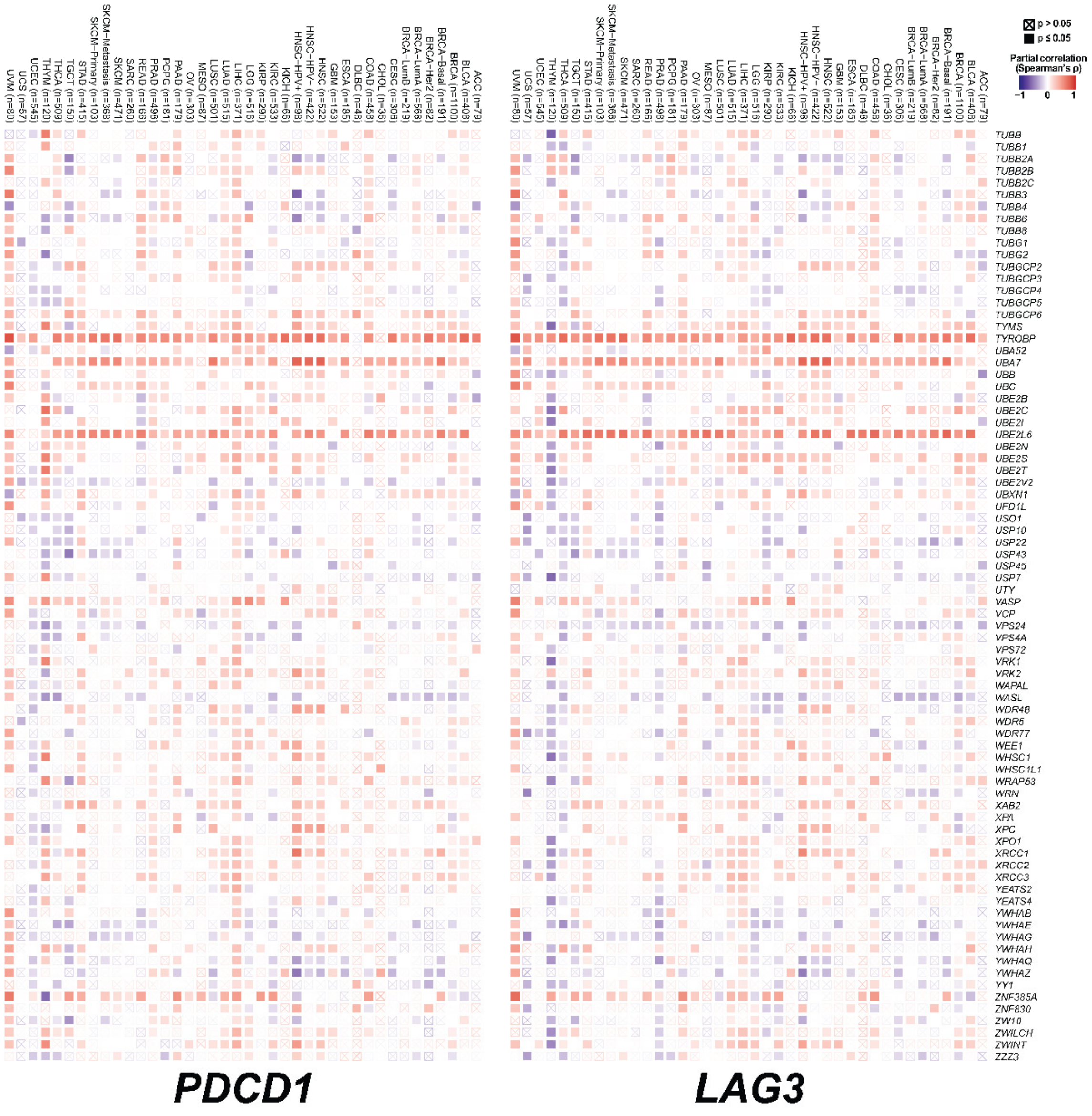


**Figure S5. Correlation of *PDCD1* and *LAG3* gene expression with genes regulating cell cycle, cell-cell communication, chromatin organization, DNA repair and DNA replication in TCGA cancers.** Heat maps show correlates of *PDCD1* and *LAG3* gene expression with the indicated genes, as shown for each cancer type (top). A partial purity-adjusted Spearman's correlation was used to test statistical significance. Red, positive correlation; blue, negative correlation. Crossed squares indicate no significant correlation (p>0.05). Specific cancer types are indicated on top, and targets on the right.

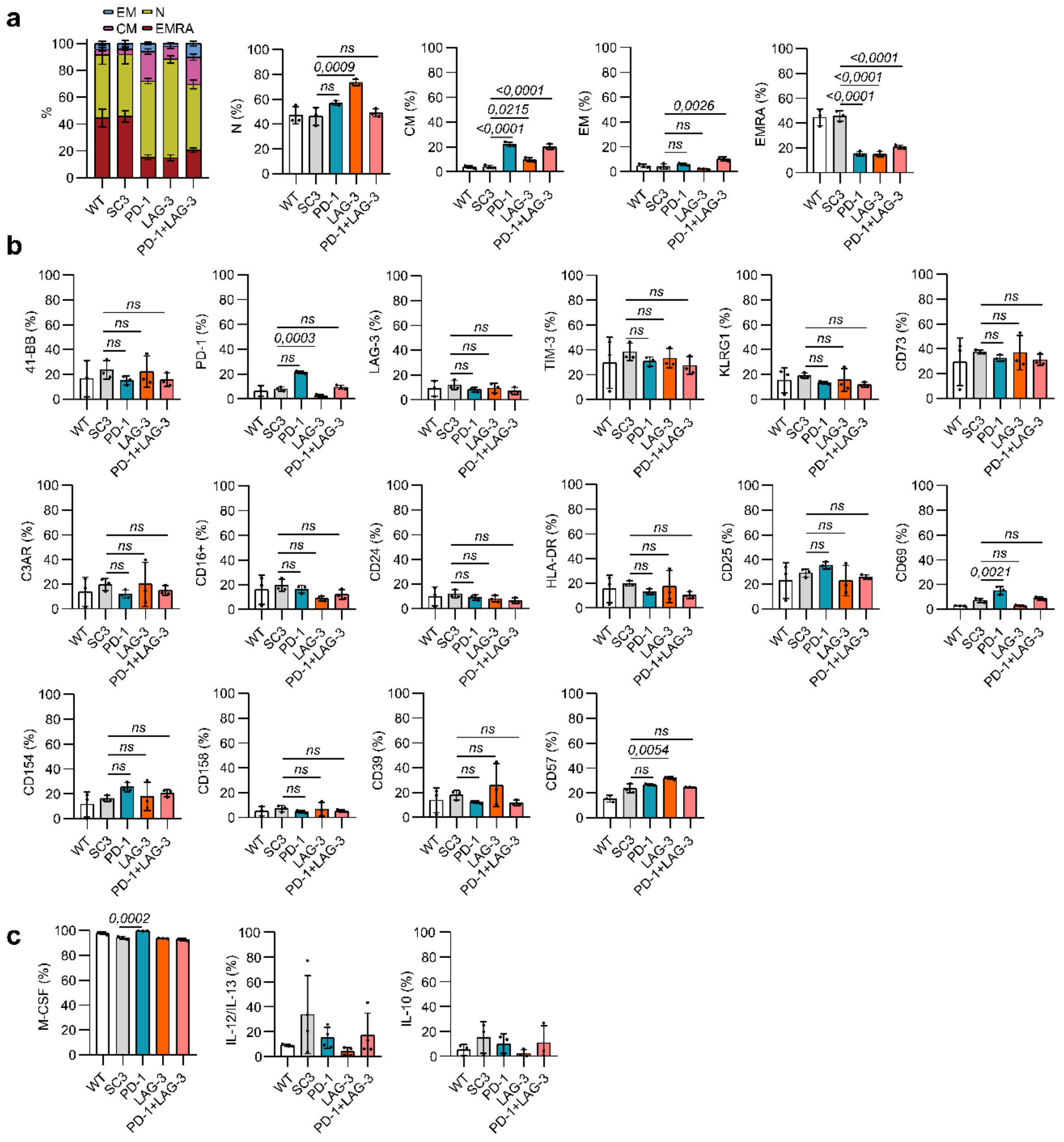

**Figure S6. Phenotypes of Jurkat CD4 T-cell lines expressing active PD-1, LAG-3 or PD-1/LAG-3 molecules. (a)** Graph bars of T-cell percentages with naïve (N), central memory (CM), effector memory (EM) and effector cell re-expressing CD45RA (EMRA) T-cells. Means from three independent experiments (n=3) are shown with standard deviations as error bars. **(b)** As in (a), but representing the percentage of T-cells expressing the indicated activation markers, immune checkpoints, exhaustion markers and other immunosuppressive markers. **(c)** Same as (a), but representing the percentage of cells expressing the indicated cytokines. WT, unmodified Jurkat T-cells; SC3, SC3-control JurkaT-cells. Two-way ANOVAs were carried out, followed by a posteriori pair-wise comparisons with Tukey's test.

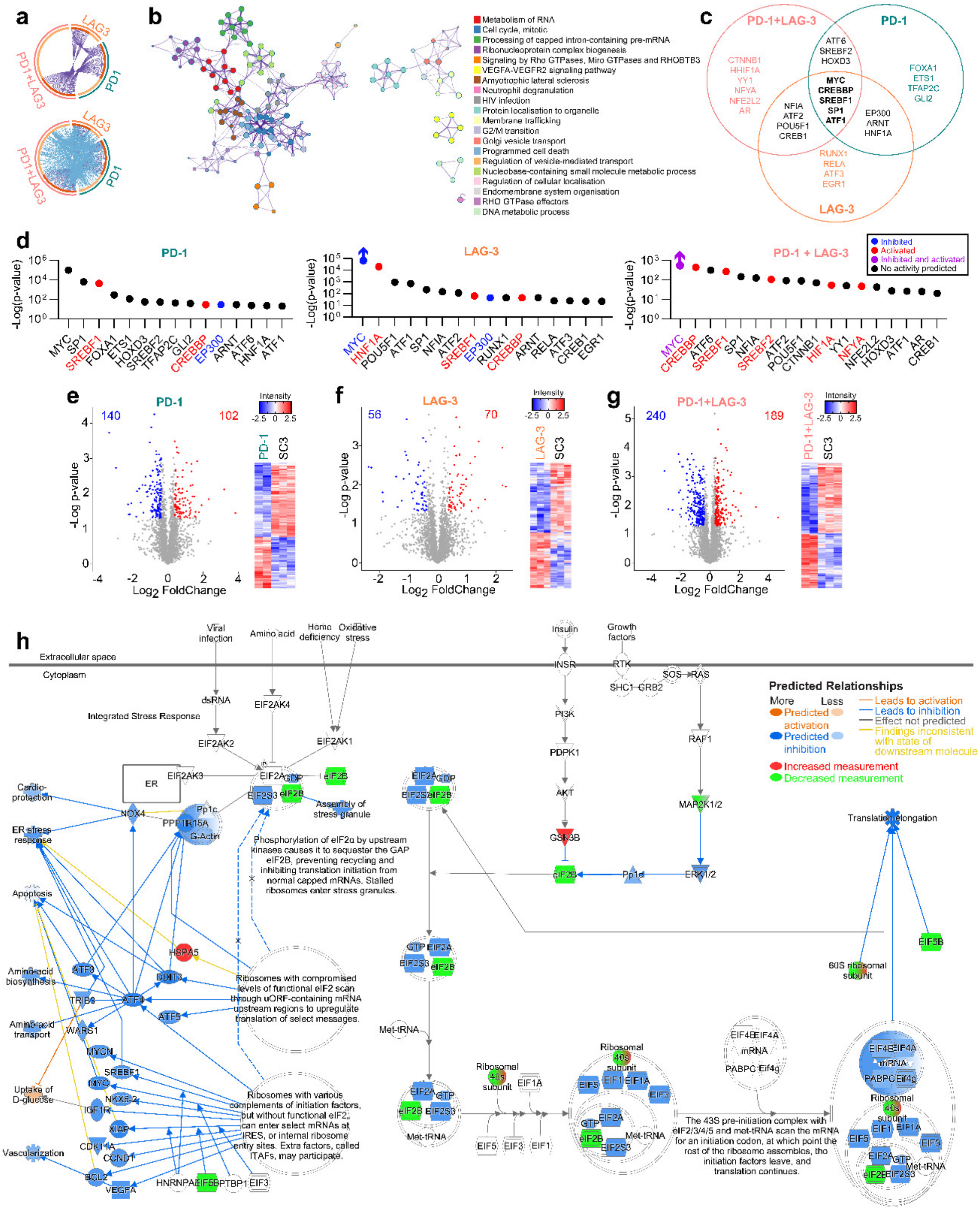

**Figure S7. Proteomes of T-cells with PD-1/LAG-3-regulated pathways. (a)** Circo plot represents the overlap from the input proteome dataset lists. Upper circle: On the outside, each arc represents the identity of each proteome. On the inside, each arc represents a list, where each gene has a spot on the arc. Dark orange colour represents the proteins that appear in multiple lists and light orange colour represents proteins that are unique to that list. Purple lines link the same protein that are shared by multiple lists. Above circle: On the outside, same as upper circle. On the inside, each arc represents a list, where each gene has a spot on the arc. Dark orange colour represents the molecules that appear in multiple lists and light orange colour represents molecules that are unique to that list. Purple lines link the same gene that are shared by multiple lists. Blue lines link the different genes where they fall into the same ontology term (the term has to statistically significantly enriched and with size no larger than 100). Blue links indicate the degree of functional overlap among the input lists. (**b**) Subset of representative terms from the full cluster converted into a network layout. Each term is represented by a circle node, where its size is proportional to the number of input molecules falling into that term, and its color represents cluster identity (i.e., nodes of the same color belong to the same cluster). Terms with a similarity score > 0.3 are linked by an edge (the thickness of the edge represents the similarity score). The network is visualized with Cytoscape (v3.1.2) with force-directed layout and with bundled edge for clarity. One term from each cluster is selected to have its term description shown as label. **(c)** Venn diagram representing the common transcription factors for the 35 common proteins on PD-1, LAG-3 and PD-1+LAG-3 proteomes. **(d)** Transcription factor predicted to regulate PD-1, LAG-3 or PD-1+LAG-3 proteomes. **(e)** **(f)** **(g)** Differential proteins for PD-1, LAG-3 or PD-1+LAG-3 vs OKT3 represented as volcano plots and heatmaps. **(h)** Constitutive activated PD-1+LAG-3 proteomic networks associated to downregulation of EIF2 signalling. The specific legends to inter-nodal relationships are described in QIAGEN IPA (**Ingenuity Pathway Analysis | QIAGEN Digital Insights**).

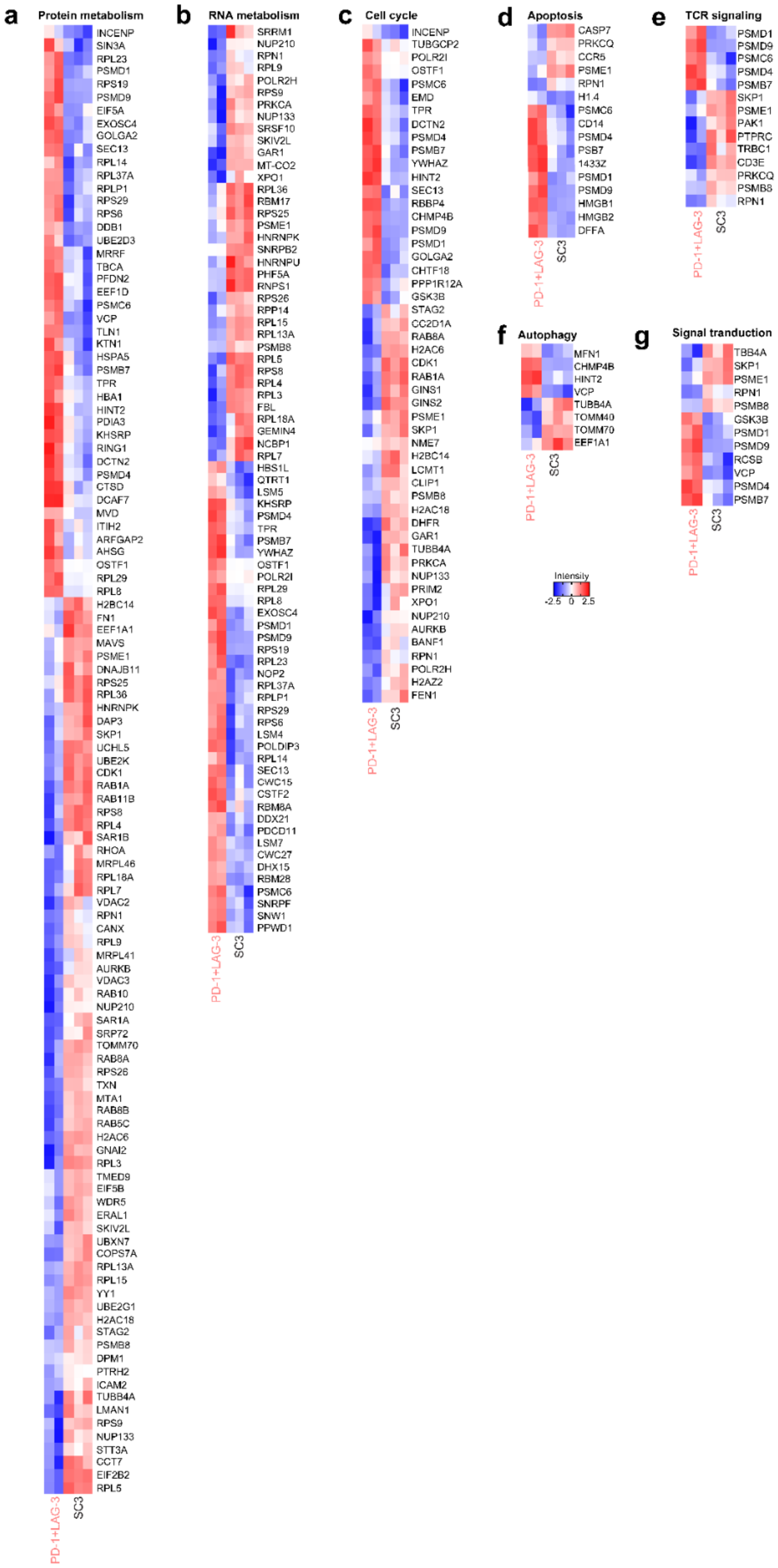


**Figure S8. Heatmaps of differential protein expression in the PD-1+LAG-3 proteomic dataset compared with the proteome of the SC3-Jurkat control cell line.** **(a)** Regulators of protein metabolism. **(b)** Regulators of RNA metabolism. **(c)** Cell cycle proteins. **(d)** Regulators of apoptosis. **(e)** TCR signalling proteins. **(f)** Autophagy regulators. **(g)** Signal transduction proteins. Red, significantly up-regulated proteins (p<0.05); Blue, significantly down-modulated proteins (p>0.05). Relevant T-cell pathways and functions are indicated on top. Specific targets are indicated on the right.

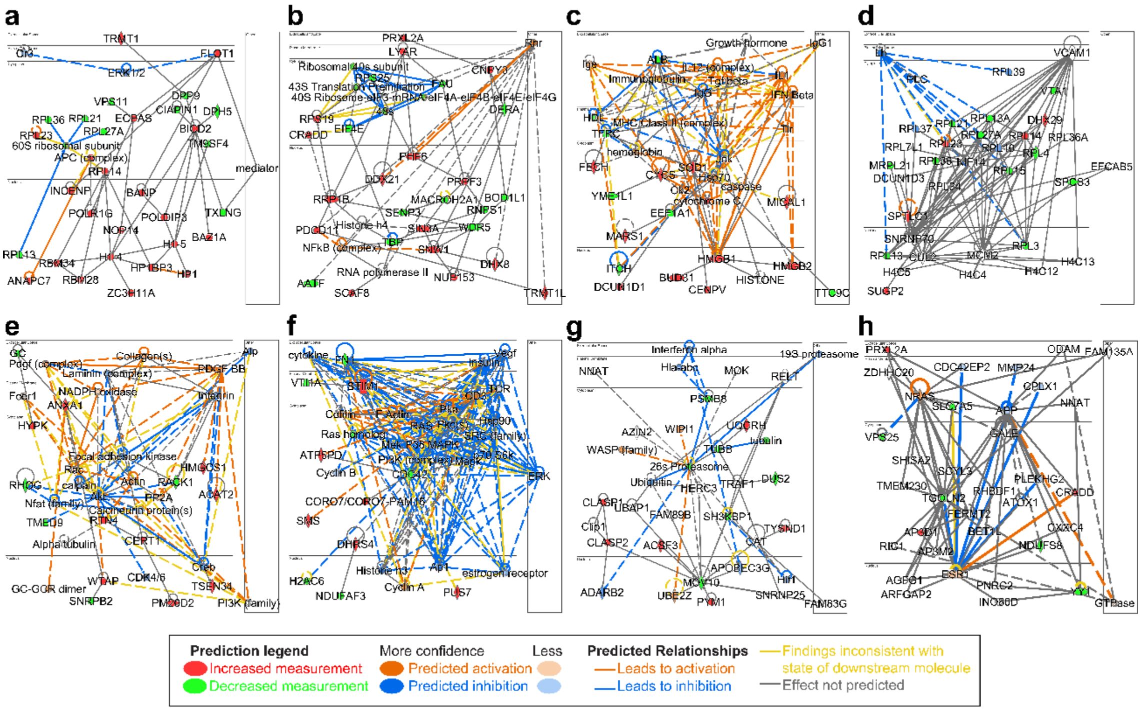


**Figure S9**. **Proteomic interactome networks associated to constitutive activation of LAG-3 in Jurkat T-cell lines, generated by IPA.** (**a**) Protein network associated with cell morphology, protein synthesis and RNA damage and repair functions; (**b**) molecular transport, RNA post-transcriptional modification, and RNA trafficking functions; **(c)** cell death and survival, free radical scavenging and organismal injury and abnormalities functions; **(d)** cellular assembly and organization, protein synthesis and RNA damage and repair functions; **(e)** cell morphology, cellular function and maintenance and cellular movement functions; **(f)** cell morphology, cellular movement, skeletal, muscular system development, and function functions; **(g)** cellular assembly and organization, cellular function and maintenance and dental disease functions; **(h)** cancer, cell-to-cell signalling and interaction, injury, and abnormalities. The specific legends to inter-nodal relationships are described in QIAGEN IPA (**Ingenuity Pathway Analysis | QIAGEN Digital Insights**).

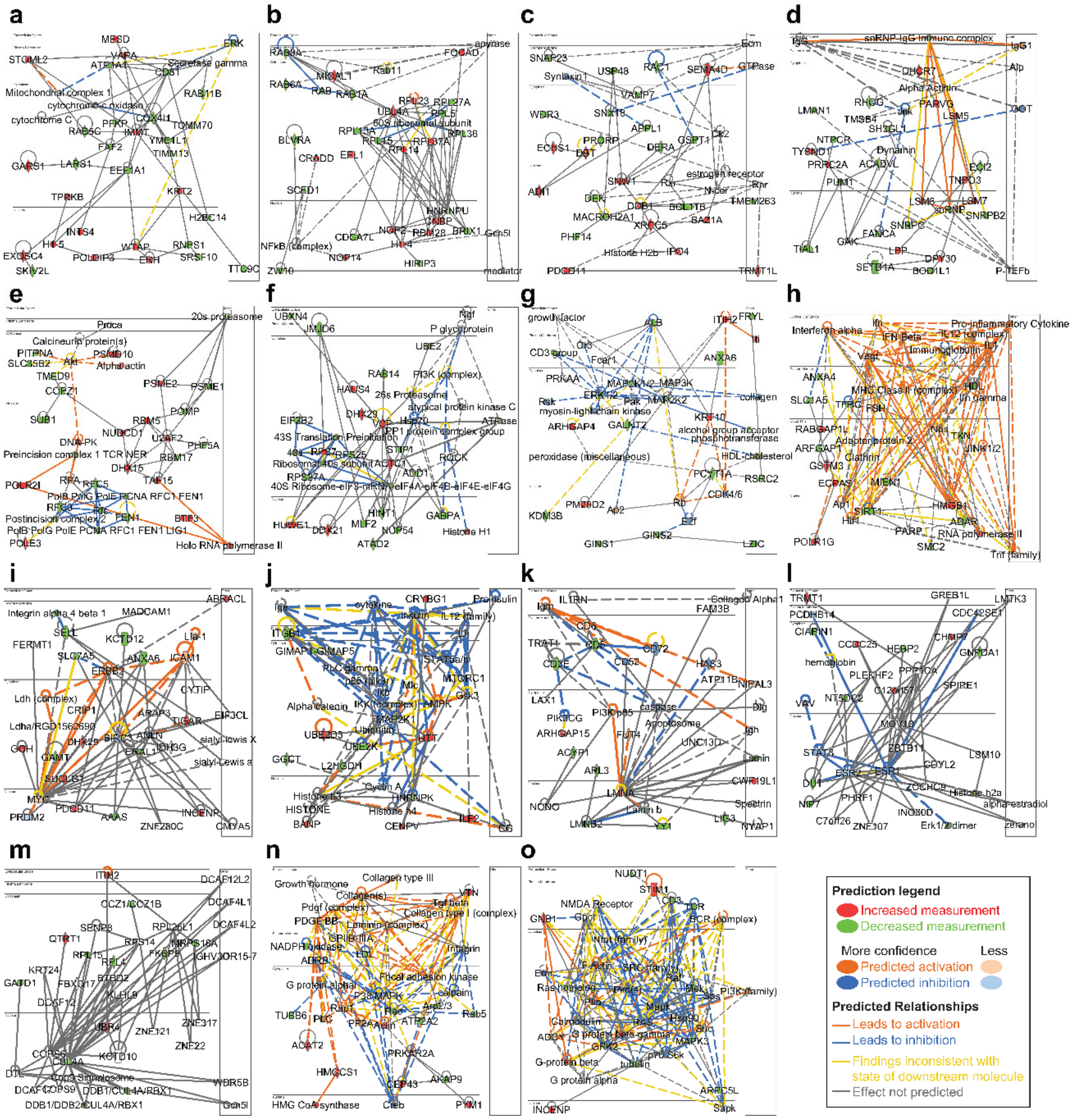


**Figure S10. Proteomic interactome networks associated to constitutive activation of PD-1 in Jurkat T-cell lines, generated by IPA.** **(a)** Protein network associated with cell-to-cell signalling and interaction, cellular assembly and organization and infectious diseases; **(b)** cancer, protein synthesis and RNA damage and repair; **(c)** cell cycle, cellular movement and nervous system development and functions; **(d)** cancer, haematological disease, and RNA post-transcriptional modification functions; **(e)** DNA replication, recombination, and repair, infectious diseases, and RNA post-transcriptional modification functions; **(f)** cancer, cell death and survival and injury and abnormalities; **(g)** cancer, cardiovascular disease, and neurological disease; **(h)** DNA replication, recombination, and repair, gene expression and nucleic acid metabolic functions; **(i)** cancer, cell-to-cell signalling and interaction and cellular compromise; **(j)** nervous system development and function, protein synthesis and tissue morphology; **(k)** haematological system development and function, humoral immune response and lymphoid tissue structure and development; **(l)** cancer, developmental disorder, and endocrine system disorders; **(m)** cellular assembly and organization, dermatological diseases and conditions and RNA damage and repair; **(n)** developmental disorder, hereditary disorder, and metabolic disease; **(o)** cellular assembly and organization, cellular compromise, and hypersensitivity response. The specific legends to inter-nodal relationships are described in QIAGEN IPA (**Ingenuity Pathway Analysis | QIAGEN Digital Insights**).

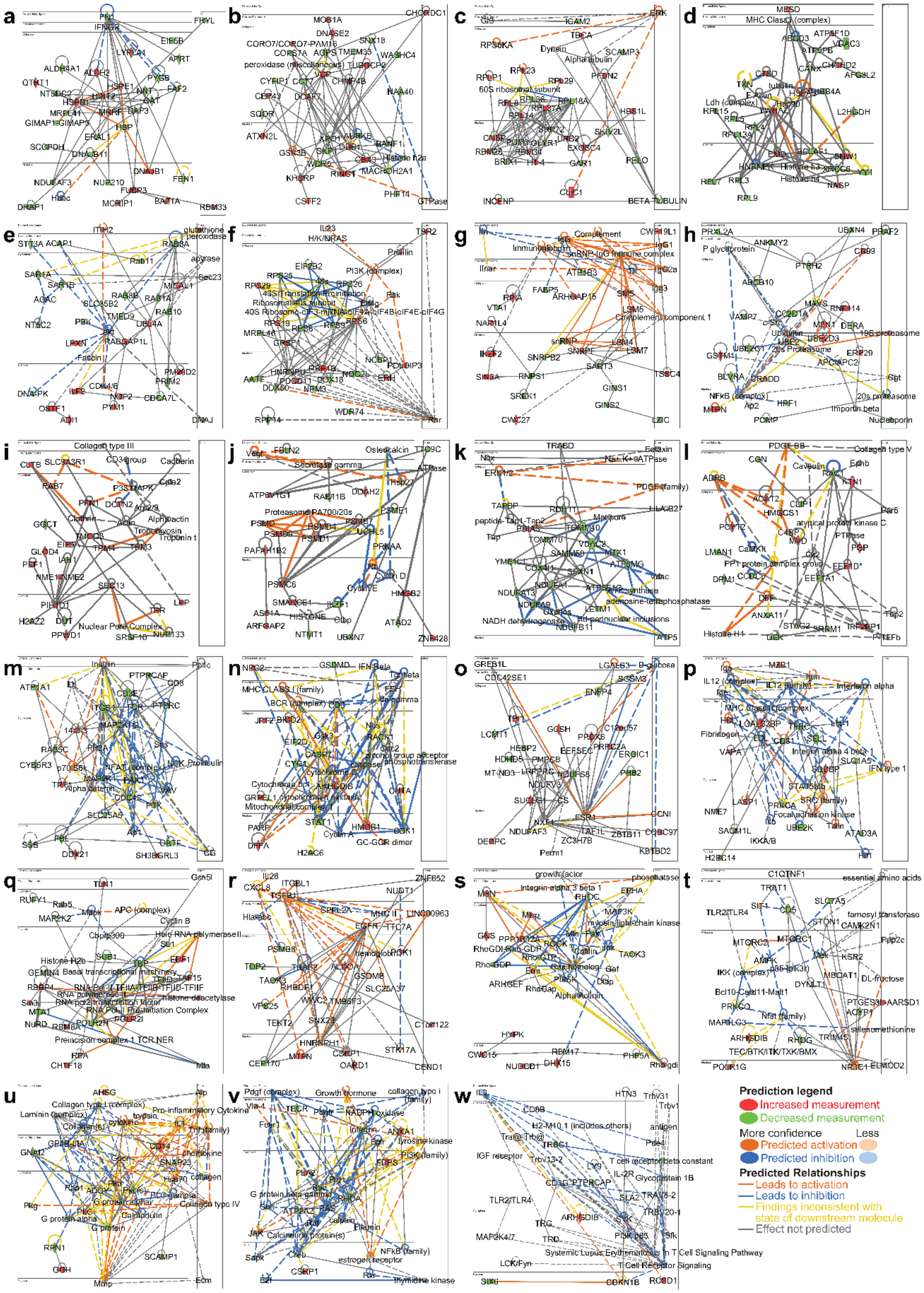


**Figure S11. Proteomic interactome networks associated to constitutive activation of PD-1 +LAG-3 in Jurkat T-cell lines, generated by IPA. (a)** Protein network associated with energy production, nucleic acid metabolism and protein synthesis; **(b)** cell cycle, cell death and survival and cellular compromise; **(c)** connective tissue disorders, protein synthesis, RNA damage and repair functions; **(d)** neurological disease, protein synthesis and RNA damage and repair; **(e)** cell morphology, energy production and immunological disease; **(f)** protein synthesis, RNA damage and repair, RNA post-transcriptional modification; **(g)** connective tissue disorders, developmental disorder, RNA post-transcriptional modification functions; **(h)** antimicrobial response, cell-to-cell signalling and interaction and inflammatory response; **(i)** cellular assembly and organization, molecular transport, and RNA trafficking; **(j)** gene expression, post-translational modification and RNA damage and repair; **(k)** cellular assembly and organization, metabolic disease and organismal injury and abnormalities; **(l)** lipid metabolism, small molecule biochemistry and vitamin and mineral metabolism functions; **(m)** cellular assembly and organization, cellular movement, and infectious diseases; **(n)** cell cycle, cell death and survival and cell-to-cell signalling and interaction; **(o)** hereditary disorder, metabolic disease, injury, and abnormalities; **(p)** cell-to-cell signalling and interaction, cellular movement and haematological system development and function. The specific legends to inter-nodal relationships are described in QIAGEN IPA (**Ingenuity Pathway Analysis | QIAGEN Digital Insights**).

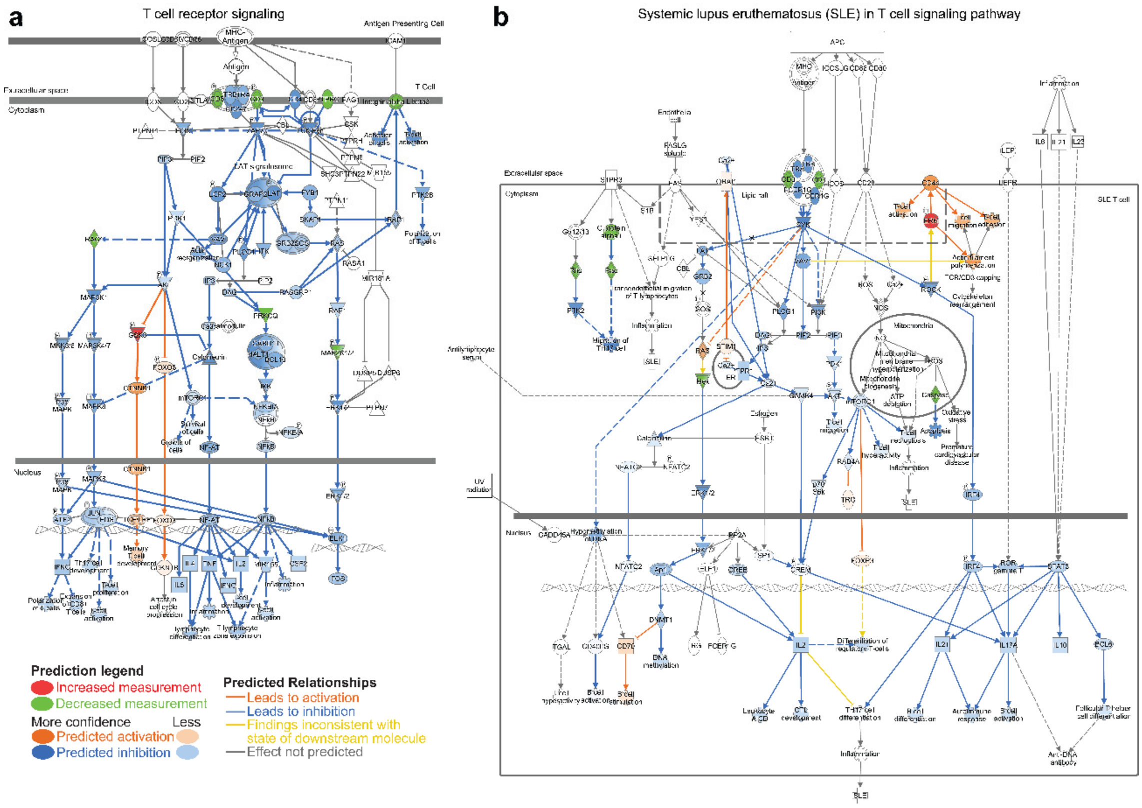


**Figure S12. IPA-reconstructed proteomic interactome networks associated to Jurkat T-cells with constitutively active *PD-1+LAG-3 signaling.* *(a)*** *Inhibition of T-cell receptor signalling.* ***(b)*** *T-cell signalling pathways associated to systemic lupus erythematosus. The specific legends to inter-nodal relationships are described in QIAGEN IPA (*[**Ingenuity Pathway Analysis | QIAGEN Digital Insights**]*).*

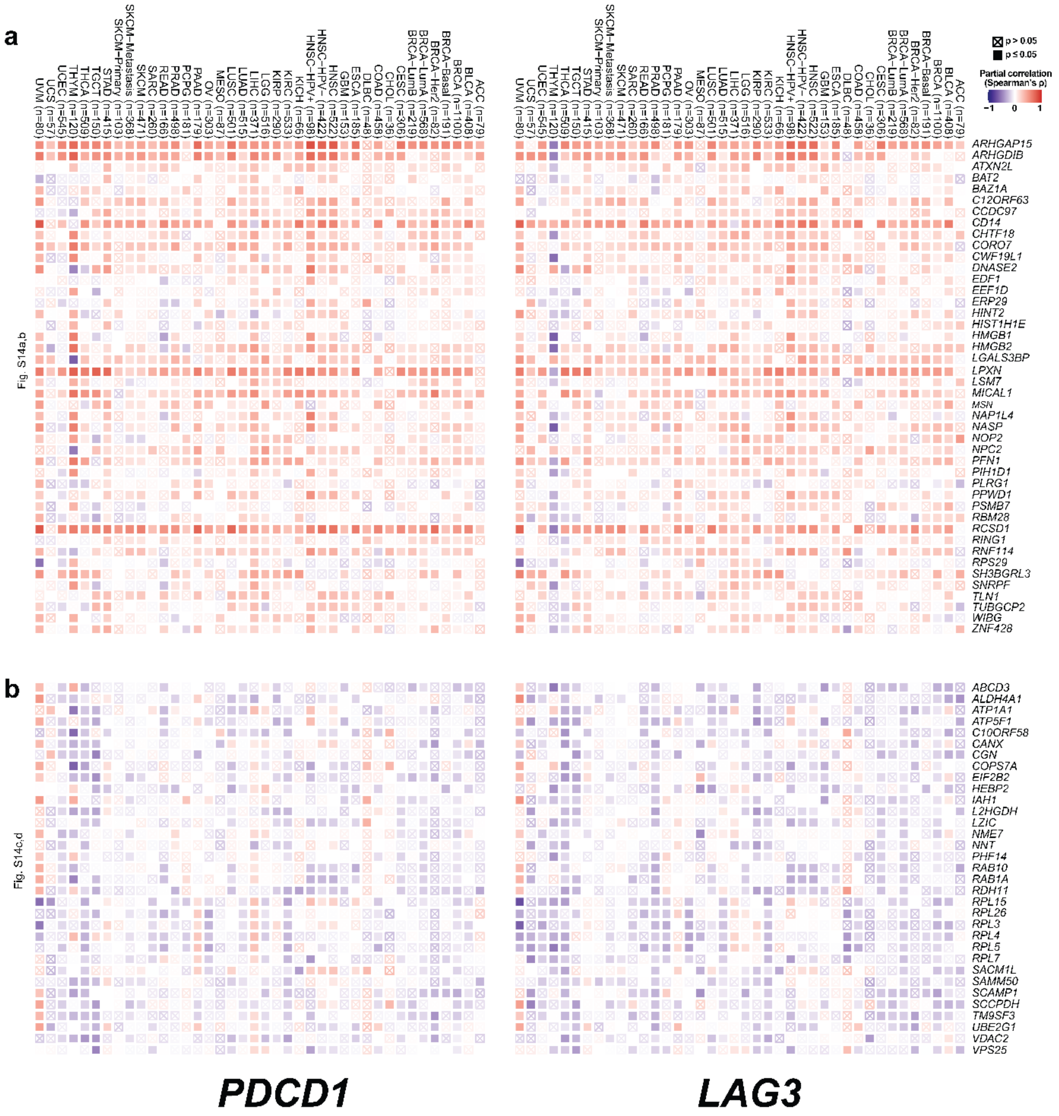


**Figure S13. Correlations of *PDCD1* and *LAG3* gene expression in TCGA cancers with selected genes corresponding to protein targets differentially expressed in the PD-1/LAG-3 T-cell proteome signature.** These targets in T-cell proteomes follow the same correlation patterns with *PDCD1* and *LAG3* gene expression in data from human tumour samples. Heat maps show correlates calculated with partial purity-adjusted Spearman's correlations to test for statistical significance. Crossed squares indicate no significant correlation (p>0.05). Red, positive correlation; Blue, negative correlation. **(a)** Significantly upregulated targets differentially expressed in the PD-1/LAG-3 proteome signature. **(b)** Significantly down-regulated targets differentially expressed in the PD-1/LAG-3 proteome signature. Specific tumour types are indicated on top. Targets are named on the right.

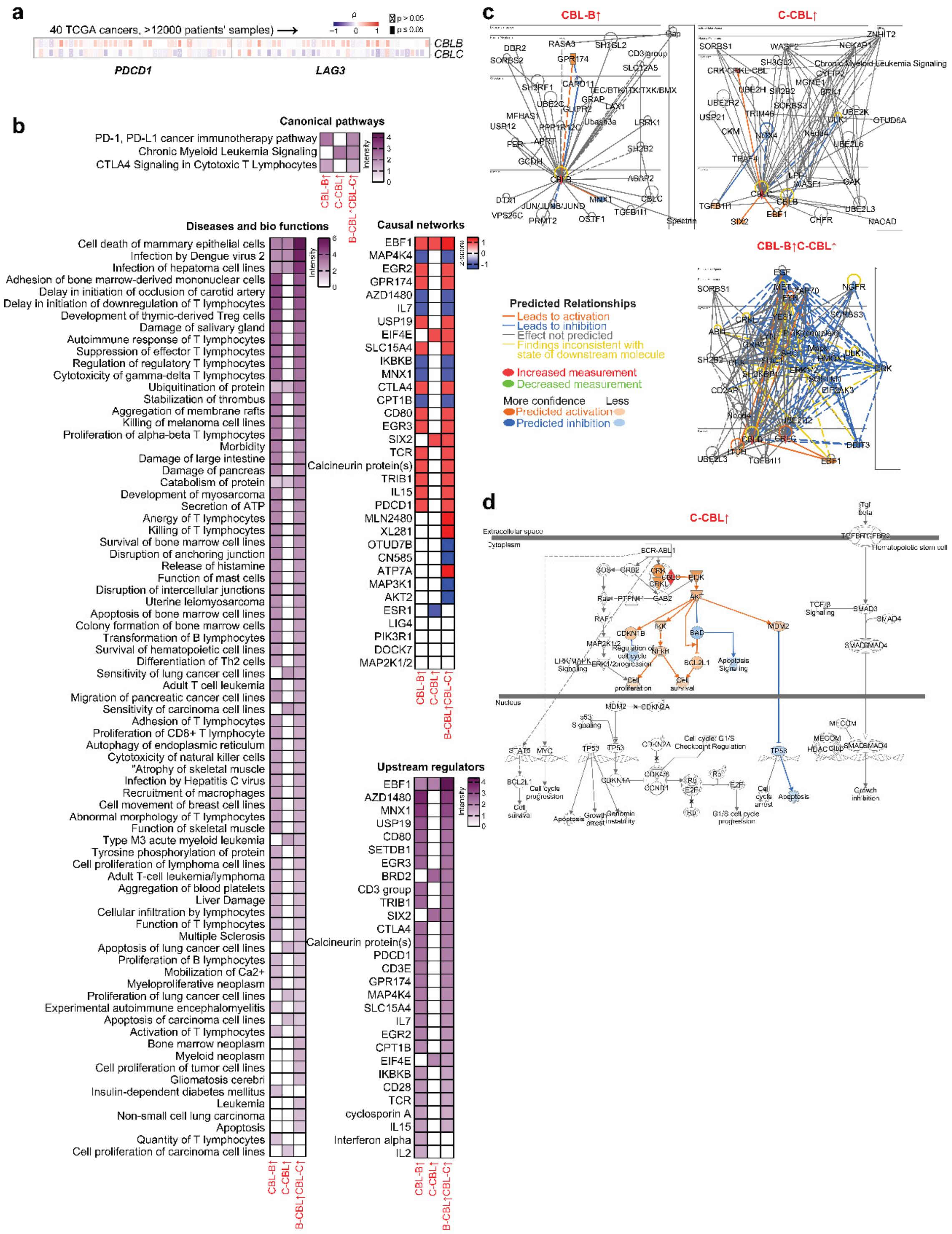

**Figure S14. Regulatory networks and causal relationships of the gene signature associated to E3 ubiquitin ligases CBL-B/C-CBL (a)** Correlation of *PDCD1* and *LAG3* gene expression with *CBLB* and *CBLC* in TCGA cancers. Red, positive correlation; Blue, negative correlation. Crossed boxes, no significant correlation by Spearman´s test. **(b)** Reconstruction of regulatory networks with up-regulated and down-regulated gene expression and causal relationships with B-CBL/CBL-C signature in T-cells, using

QIAGEN IPA algorithms. Data from curated publicly available datasets of RNA-seq, small RNA-seq, metabolomics, proteomics, microarrays including miRNA and SNP, and small-scale experiments were used. Enrichment intensities of the indicated canonical pathways are represented as a function of up-regulation of CBL-B, C-CBL or both as shown. The identified causal networks as indicated in the figure are correlated with z-scores to up- or down-regulation of B-CBL, CBL-C or both as shown. The enrichment intensities for the indicated upstream regulators are represented as a function of up-regulation of CBL-B, C-CBL or both as shown. The enrichment intensities for the indicated diseases and biofunctions as identified by IPA algorithms are as a function of up-regulation of CBL-B, C-CBL or both as shown. **(c)** Reconstructed regulatory interactomes and networks associated to the activation of CBL-B, C-CBL or both. **(d)** Reconstructed regulatory interactome of chronic myeloid leukemia signalling networks associated to the activation of CBL-C. Specific legends to inter-nodal relationships are described in QIAGEN IPA (**Ingenuity Pathway Analysis | QIAGEN Digital Insights**).
